# Prediction Model for Detection of Sporadic Pancreatic Cancer (PRO-TECT) in a Population-Based Cohort Using Machine Learning and Further Validation in a Prospective Study

**DOI:** 10.1101/2022.02.14.22270946

**Authors:** Wansu Chen, Yichen Zhou, Fagen Xie, Rebecca K. Butler, Christie Y. Jeon, Tiffany Q. Luong, Yu-Chen Lin, Eva Lustigova, Joseph R. Pisegna, Sungjin Kim, Bechien U. Wu

## Abstract

**OBJECTIVES:** There is currently no widely accepted approach to screening for pancreatic cancer (PC). We aimed to develop and validate a risk prediction model for PC across two health systems using electronic health records (EHR).

**METHODS:** This retrospective cohort study consisted of patients 50-84 years of age meeting utilization criteria in 2008-2017 at Kaiser Permanente Southern California (KPSC, model training, internal validation) and the Veterans Affairs (VA, external validation). ‘Random survival forests’ models were built to identify the most relevant predictors from >500 variables and to predict PC within 18 months of cohort entry. A prospective study was then conducted in KPSC to assess feasibility of the model for real-time implementation.

**RESULTS:** The KPSC cohort consisted of 1.8 million patients (mean age 61.6) with 1,792 PC cases. The estimated 18-month incidence rate of PC was 0.77 (95% CI 0.73-0.80)/1,000 person-years. The three models containing age, abdominal pain, weight change and two laboratory biomarkers (ALT change/HgA1c, rate of ALT change/HgA1c, or rate of ALT change/rate of HgA1c change) had comparable discrimination and calibration measures (c-index: mean=0.77, SD=0.01-0.02; calibration test: p-value 0.2-0.4, SD 0.2-0.3). The VA validation cohort consisted of 2.6 million patients (mean age 66.1) with an 18-month incidence rate of 1.27 (1.23-1.30). A total of 606 patients were screened in the prospective pilot study at KPSC with 9 patients (1.5%) diagnosed with a pancreatic or biliary cancer.

**CONCLUSIONS:** Using widely available parameters in EHR, we developed a population-based parsimonious model for early detection of sporadic PC suitable for real-time application.

**Study Highlights:** *What Is Known:*

- Patients with pancreatic cancer are often diagnosed at late stages.
- Early detection is needed to impact the natural history of disease progression and improve patient survival.

*What Is New Here:*

- Machine-learning was used to develop a population-based model for early detection of pancreatic cancer. The model was internally and externally validated in cohorts of 1.8 million and 2.6 million individuals, respectively.
- Calibration was excellent in prospective pilot testing for detection of pancreatic malignancy.

## INTRODUCTION

Pancreatic cancer is the third leading cause of cancer deaths with 48,220 estimated deaths in 2021 in the US.^1^ Because of the lack of an early detection strategy, majority of patients (50-55%) have metastases at distant sites at the time of diagnosis.^2,3^ Once diagnosed, the average 5-year survival is only 10.8%.^1^ Accounting for 90% of all pancreatic cancer cases, pancreatic ductal adenocarcinoma (PDAC) is by far the most common form of pancreatic cancer, and also the most lethal.

Due to the low incidence of pancreatic cancer in the general population (13.2 per 100,000 person-years),^1^ widespread population-based screening is not currently recommended by the United States Preventative Services Task Force.^4^ Therefore, alternative approaches to early detection are needed in order to substantially impact the natural history of this disease and improve survival for patients.

The emergence of comprehensive EHR and maturation of machine learning offers an opportunity to enhance efforts in early detection in pancreatic cancer. To date, efforts to develop clinical prediction models in pancreatic cancer have focused on specific populations such as those with new-onset diabetes^5-7^or within the confines of a case-control study.^8,9^ Efforts to target the high-risk patients in the general population are sparse.^10^ There is a critical need for novel risk stratification tools which are both sensitive and specific for rapid identification of patients at increased risk of developing pancreatic cancer.

The aim of the present study was to develop and validate a clinical prediction model for risk of PDAC across several large health systems. Specifically, we sought to apply machine-learning combined with a comprehensive approach to data in EHR to predict the risk of sporadic PDAC.

## METHODS

### Study Design and Setting

We conducted a retrospective cohort study utilizing multi-ethnic health plan enrollees of Kaiser Permanente Southern California (KPSC), a large integrated healthcare system that provides comprehensive healthcare services for >4.7 million enrollees across 15 medical centers and 235 medical offices. Model training and internal validation were conducted based on EHR data. The demographics and socioeconomic status of KPSC health plan enrollees are comparable to those of residents in the Southern California region.^11^ The internally validated models were externally tested using EHR of Veterans Affairs (VA).^12^ The study protocol was approved by the KPSC’s institutional Review Board.

### Study Population

#### Model training and internal validation

Patients 50-84 years of age and had ≥1 clinic-based visit (index visit) within a KPSC facility in 2008-2017 were identified. Patients who had history of pancreatic cancer, or not continuously enrolled in the KPSC health plan in the past 12 months (gaps 45 days or less were allowed) were excluded. The requirement of continuous enrollment allowed adequate data to define study variables. For patients with multiple qualifying index visits, we selected one randomly as the index visit. The corresponding visit date was referred to as the index date (t_0_). Follow-up started on t_0_ and ended with the earliest of the following events: disenrollment from the health plan, end of the study (December 31, 2018), reached the maximum length of follow-up (18 months), non-PDAC related death, or PDAC diagnosis or death (outcome). A minimum of 30 days of follow-up is required.

#### Model testing

Veterans 50-84 years of age who had >1 outpatient visit (index visit) within a VA facility in 2008-2017 and another clinic-based visit within the 12 months prior to the index date were identified. Patients who had history of pancreatic cancer were excluded. The same follow-up rules mentioned above were applied to the VA cohort except for “disenrollment from the health plan”.

### Outcome Identification

The study outcome was PDAC diagnosis or death with pancreatic cancer in the 18 months after the index date. For the KPSC cohort, PDAC was identified from the Cancer Registry by using the Tenth Revision of International Classification of Diseases, Clinical Modification (ICD-10-CM) code C25.x and histology codes (eTable 1). Pancreatic cancer deaths were derived from the linkage with the California State Death Master files and identified using ICD-10-CM codes C25.x.^13^ For the VA cohort, cases of PDAC were similarly identified through an internal VA Central Cancer Registry, and PDAC deaths identified through the VA Mortality Data Repository, which integrates vital status data from the National Death Index (NDI), VA, and DoD administrative records.

### Patient Demographic and Clinical Features at Baseline

A complete list of extracted and derived features for the KPSC cohort is shown in eTable 2. Except for demographic variables, values within each time interval (0-6 months, 7-12 months, 1-2 years and >2 years) were generated. Definitions of the derived variables were described in eTable3. Since the VA dataset was solely used for testing purposes, only limited number of features were extracted (Table 1).

**Table 1.**
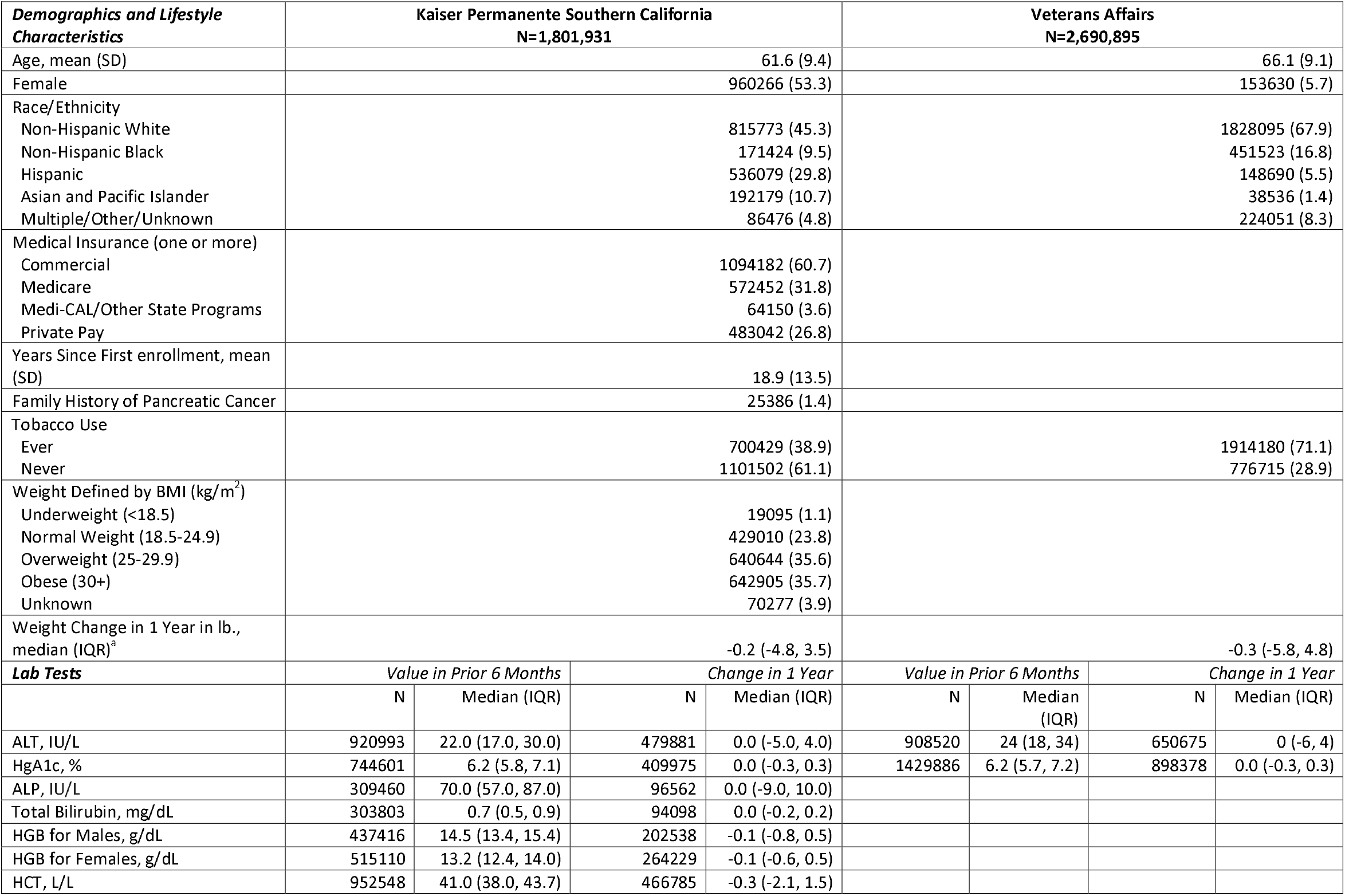
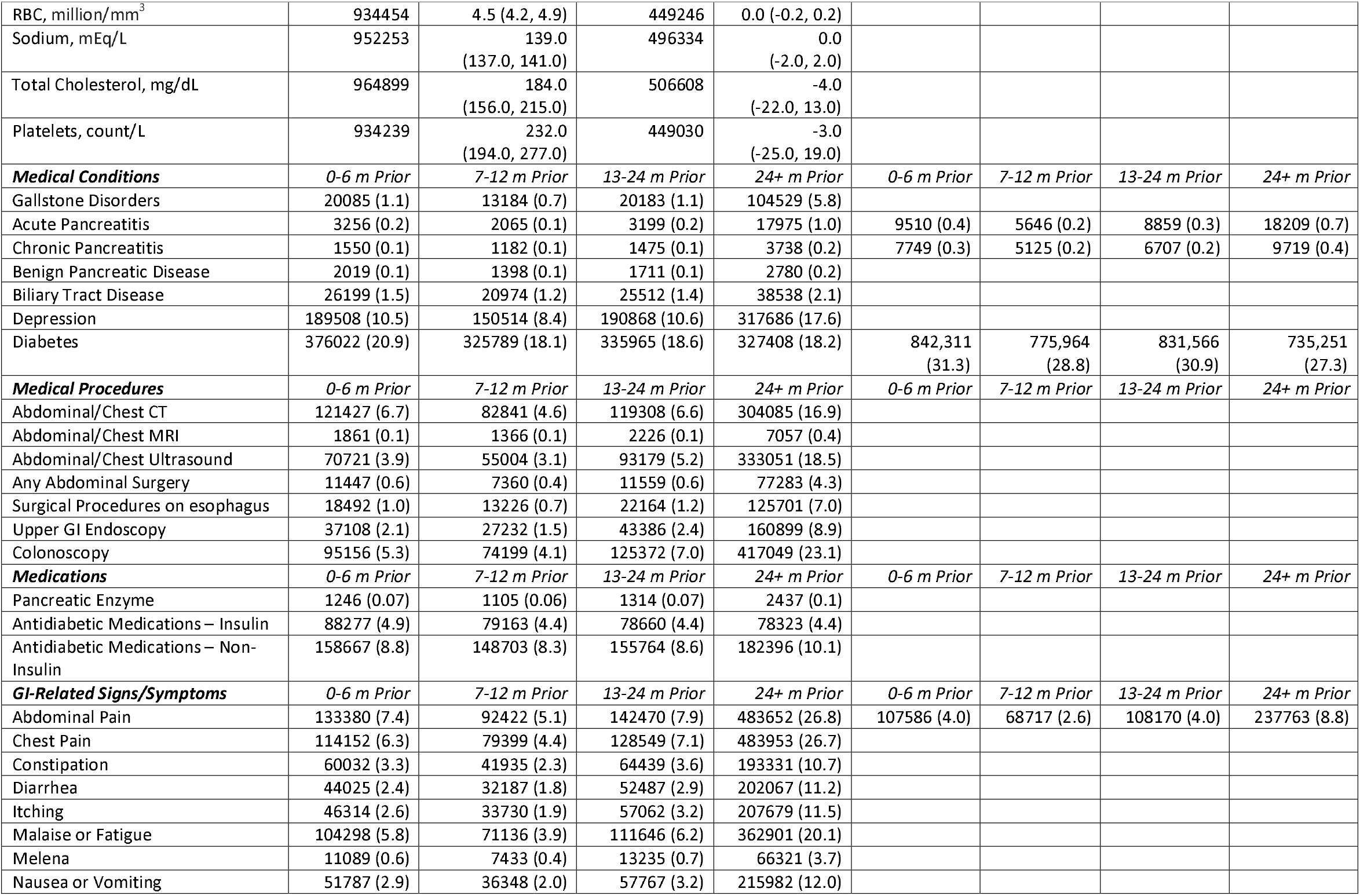
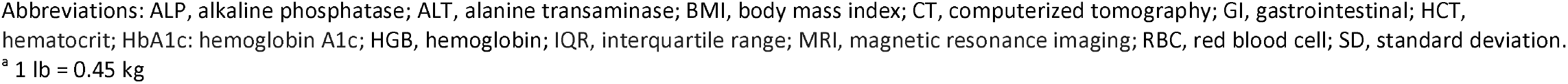
Characteristics of study subjects at baseline, n (%) unless otherwise stated.

Missing values were imputed^14^ if the frequency of missing was <60%. We used predictive mean matching method^15^ with k=5. Laboratory measures with ≥60% missingness or change/change rate measures with ≥80% missingness were not included in the model development process. Ten imputed datasets were generated.

### Model Training, Validation and Testing

To overcome the limitations of regression-based models that are traditionally used for analysis of time-to-event data, we applied ‘random survival forests’ (RSF), a nonparametric machine learning method,^16-18^ to pre-select features and train/validate models. First, we iteratively preselected features based on the average minimum depth (eTable 4) and used those features to develop and validate risk prediction models based on 5-fold cross validation.^19^ Age was forced into the model. Preselected features were added incrementally to identify the feature that yielded the maximum improvement of c-index. This process continued until the c-index increased <0.005. Of the 50 models derived from the 50 training datasets (10 imputation datasets x 5-fold cross validation), the three that appeared the most often were selected as the winning models.

Algorithms of the winning models were applied to the corresponding KPSC validation datasets. By design, the KPSC validation datasets did not include any observations of the KPSC training datasets from which the winning models were developed. One winning model was first directly applied to VA imputed datasets, and subsequently recalibrated to achieve better performance.

### Performance Measures

The discriminative power for each of the winning models was evaluated by c-index, a concordance measure, averaged across all the relevant validation datasets for cohort members. Calibration was assessed by calibration plots with five risk groups (<50th, 50–74th, 75–89th, 90–94th, 95–100th percentiles).^20^ Greenwood-Nam-D’Agostino (GND) calibration test was also performed to assess goodness-of-fit.

We estimated sensitivity, specificity, positive predictive value (PPV), and relative increase in risk in comparison to that of the entire cohort at various levels of risk thresholds. For this analysis we restricted the patients to those with complete follow up or developed PDAC in 18 months. The results were averaged across the validation datasets for each winning model.

### Early Detection Model

To facilitate earlier detection of PDAC by ≥90 days, we also established a cohort which included patients identified in the main cohort who had ≥90 days of cancer-free follow up. The same model training and validation methods mentioned above were applied.

### Prospective pilot study

We conducted a subsequent prospective study from February to December 2021 to evaluate feasibility of real-time implementation of the final prediction model. We aimed to assess the calibration of the model (frequency of observed vs. expected cancer). This study was approved as a separate protocol by the KPSC Institutional Review Board. We prospectively ran a final algorithm on a bi-weekly basis to identify patients aged 50-84 without a prior history of pancreatic cancer whose predicted risk of pancreatic cancer is ≥1%. All algorithm-identified patients were manually reviewed. Patients with findings suspicious for a pancreatic cancer on cross-sectional imaging obtained through routine clinical care up to 3 months prior to the risk identification date (the date when the algorithm was applied to EHR) were monitored. The diagnosis was based on either histology where available or presumptive clinical diagnosis as documented in patients’ EHR.

### Statistical Analysis

Analyses were performed using SAS (Version 9.4 for Unix; SAS Institute, Cary, NC) or R Version 3.6.0 (R Foundation, Vienna, Austria).

## RESULTS

### Characteristics of the study cohorts

1.8 million KPSC patients were eligible (eFigure 1), of which 53.3% were females, 45.3% were white, 29.8% were Hispanic, 9.5% were African American and 10.7% were Asian and Pacific Islanders (Table 1). The majority (60.7%) used commercial insurance, and slightly under one-third (31.8%) were on Medicare. On average, the KPSC patients were 61.6 years of age, with average membership length of 18.9 years. 35.7% of the patients were obese and additional 35.6% were overweight. Diabetes was common (about 20%), while acute and chronic pancreatitis were rare (<1%).

The 2.6 million eligible veterans were predominantly male (94.3%), white (67.9%) and African American (16.8%) and were older (66.1 years of age) than the KPSC cohort. Smoking, diabetes, acute and chronic pancreatitis were more prevalent in the VA cohort compared to those of the KPSC cohort. ALT and HbA1c at baseline appeared comparable between the two cohorts.

### Incidence of PDAC

Table 2 displays the follow-up time in years, number, incidence rate (IR) of PDAC, and time to PDAC for all patients and for subgroups of patients defined by important features. 1,792 KPSC patients developed PDAC within 18 months of follow-up (IR=0.77, 95% CI 0.73-0.80/1,000 person-years (PY) (Table 2). A total of 4,582 patients in the VA cohort developed PDAC (IR 1.27 (1.23-1.30)). In the VA cohort, abdominal pain in the 6 months prior to t_0_ increased the IR to 4.35 (4.01-4.70). Time to PDAC appeared to be longer for the VA cohort (median 233 days, IQR 116-370 days) compared to that of the KPSC cohort (median 205 days, IQR 91-358 days). The distributions of cancer stage (I-IV) were comparable between the two cohorts (eTable 5), if the higher frequency of missingness in the VA cohort is ignored.

**Table 2.**
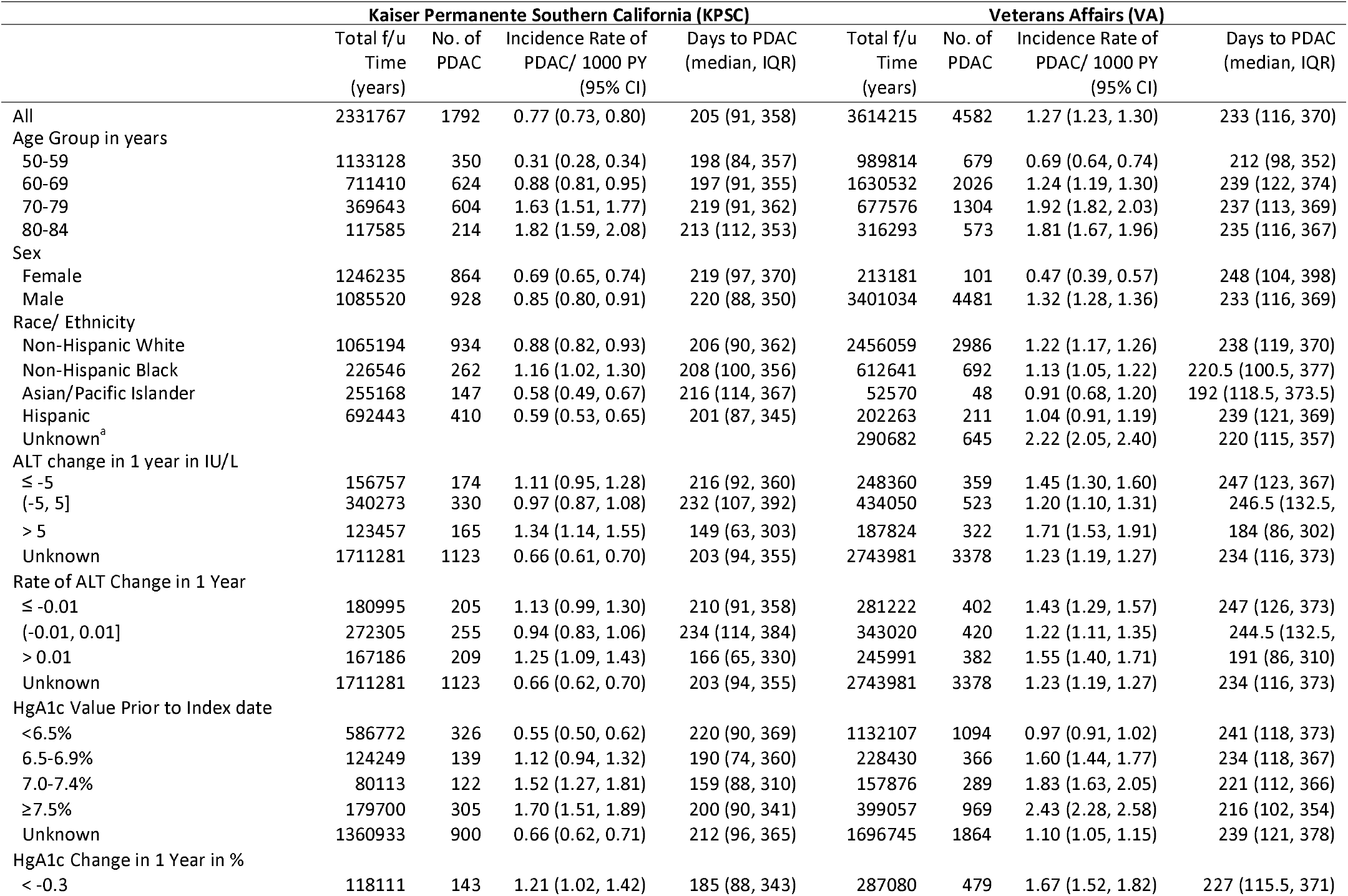
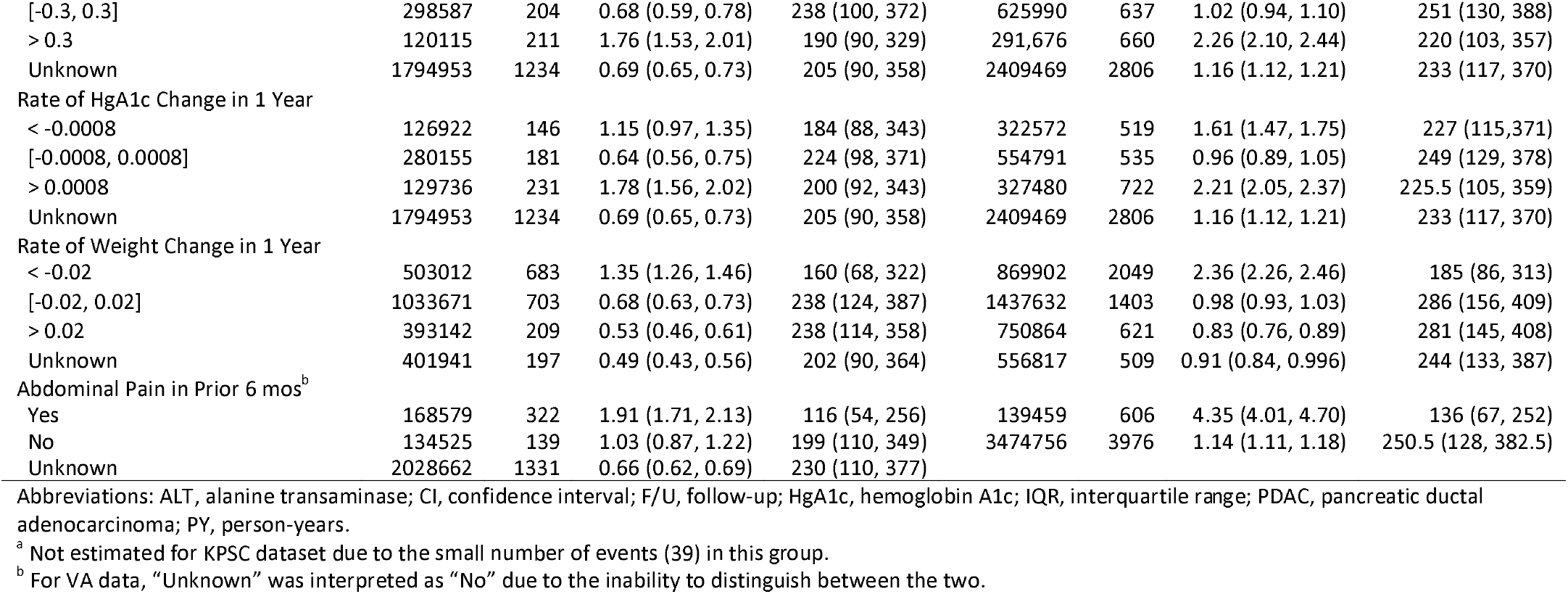
Total follow-up (f/u) time, number, and incidence rate of PDAC per 1,000 person-years (PY) and 95% CI.

|  | Kaiser Permanente Southern California (KPSC) |  |  |  | Veterans Affairs (VA) |  |  |  |
| --- | --- | --- | --- | --- | --- | --- | --- | --- |
|  | Total f/u Time (years) | No. of PDAC | Incidence Rate of PDAC/ 1000 PY (95% CI) | Days to PDAC (median, IQR) | Total f/u Time (years) | No. of PDAC | Incidence Rate of PDAC/ 1000 PY (95% CI) | Days to PDAC (median, IQR) |
| All | 2331767 | 1792 | 0.77 (0.73, 0.80) | 205 (91, 358) | 3614215 | 4582 | 1.27 (1.23, 1.30) | 233 (116, 370) |
| Age Group in years |  |  |  |  |  |  |  |  |
| 50-59 | 1133128 | 350 | 0.31 (0.28, 0.34) | 198 (84, 357) | 989814 | 679 | 0.69 (0.64, 0.74) | 212 (98, 352) |
| 60-69 | 711410 | 624 | 0.88 (0.81, 0.95) | 197 (91, 355) | 1630532 | 2026 | 1.24 (1.19, 1.30) | 239 (122, 374) |
| 70-79 | 369643 | 604 | 1.63 (1.51, 1.77) | 219 (91, 362) | 677576 | 1304 | 1.92 (1.82, 2.03) | 237 (113, 369) |
| 80-84 | 117585 | 214 | 1.82 (1.59, 2.08) | 213 (112, 353) | 316293 | 573 | 1.81 (1.67, 1.96) | 235 (116, 367) |
| Sex |  |  |  |  |  |  |  |  |
| Female | 1246235 | 864 | 0.69 (0.65, 0.74) | 219 (97, 370) | 213181 | 101 | 0.47 (0.39, 0.57) | 248 (104, 398) |
| Male | 1085520 | 928 | 0.85 (0.80, 0.91) | 220 (88, 350) | 3401034 | 4481 | 1.32 (1.28, 1.36) | 233 (116, 369) |
| Race/ Ethnicity |  |  |  |  |  |  |  |  |
| Non-Hispanic White | 1065194 | 934 | 0.88 (0.82, 0.93) | 206 (90, 362) | 2456059 | 2986 | 1.22 (1.17, 1.26) | 238 (119, 370) |
| Non-Hispanic Black | 226546 | 262 | 1.16 (1.02, 1.30) | 208 (100, 356) | 612641 | 692 | 1.13 (1.05, 1.22) | 220.5 (100.5, 377) |
| Asian/Pacific Islander | 255168 | 147 | 0.58 (0.49, 0.67) | 216 (114, 367) | 52570 | 48 | 0.91 (0.68, 1.20) | 192 (118.5, 373.5) |
| Hispanic | 692443 | 410 | 0.59 (0.53, 0.65) | 201 (87, 345) | 202263 | 211 | 1.04 (0.91, 1.19) | 239 (121, 369) |
| Unknown<sup>a</sup> |  |  |  |  | 290682 | 645 | 2.22 (2.05, 2.40) | 220 (115, 357) |
| ALT change in 1 year in IU/L |  |  |  |  |  |  |  |  |
| ≤ -5 | 156757 | 174 | 1.11 (0.95, 1.28) | 216 (92, 360) | 248360 | 359 | 1.45 (1.30, 1.60) | 247 (123, 367) |
| (-5, 5] | 340273 | 330 | 0.97 (0.87, 1.08) | 232 (107, 392) | 434050 | 523 | 1.20 (1.10, 1.31) | 246.5 (132.5, |
| > 5 | 123457 | 165 | 1.34 (1.14, 1.55) | 149 (63, 303) | 187824 | 322 | 1.71 (1.53, 1.91) | 184 (86, 302) |
| Unknown | 1711281 | 1123 | 0.66 (0.61, 0.70) | 203 (94, 355) | 2743981 | 3378 | 1.23 (1.19, 1.27) | 234 (116, 373) |
| Rate of ALT Change in 1 Year |  |  |  |  |  |  |  |  |
| ≤ -0.01 | 180995 | 205 | 1.13 (0.99, 1.30) | 210 (91, 358) | 281222 | 402 | 1.43 (1.29, 1.57) | 247 (126, 373) |
| (-0.01, 0.01] | 272305 | 255 | 0.94 (0.83, 1.06) | 234 (114, 384) | 343020 | 420 | 1.22 (1.11, 1.35) | 244.5 (132.5, |
| > 0.01 | 167186 | 209 | 1.25 (1.09, 1.43) | 166 (65, 330) | 245991 | 382 | 1.55 (1.40, 1.71) | 191 (86, 310) |
| Unknown | 1711281 | 1123 | 0.66 (0.62, 0.70) | 203 (94, 355) | 2743981 | 3378 | 1.23 (1.19, 1.27) | 234 (116, 373) |
| HgA1c Value Prior to Index date |  |  |  |  |  |  |  |  |
| <6.5% | 586772 | 326 | 0.55 (0.50, 0.62) | 220 (90, 369) | 1132107 | 1094 | 0.97 (0.91, 1.02) | 241 (118, 373) |
| 6.5-6.9% | 124249 | 139 | 1.12 (0.94, 1.32) | 190 (74, 360) | 228430 | 366 | 1.60 (1.44, 1.77) | 234 (118, 367) |
| 7.0-7.4% | 80113 | 122 | 1.52 (1.27, 1.81) | 159 (88, 310) | 157876 | 289 | 1.83 (1.63, 2.05) | 221 (112, 366) |
| ≥7.5% | 179700 | 305 | 1.70 (1.51, 1.89) | 200 (90, 341) | 399057 | 969 | 2.43 (2.28, 2.58) | 216 (102, 354) |
| Unknown | 1360933 | 900 | 0.66 (0.62, 0.71) | 212 (96, 365) | 1696745 | 1864 | 1.10 (1.05, 1.15) | 239 (121, 378) |
| HgA1c Change in 1 Year in % |  |  |  |  |  |  |  |  |
| < -0.3 | 118111 | 143 | 1.21 (1.02, 1.42) | 185 (88, 343) | 287080 | 479 | 1.67 (1.52, 1.82) | 227 (115.5, 371) |
| [-0.3, 0.3] | 298587 | 204 | 0.68 (0.59, 0.78) | 238 (100, 372) | 625990 | 637 | 1.02 (0.94, 1.10) | 251 (130, 388) |
| > 0.3 | 120115 | 211 | 1.76 (1.53, 2.01) | 190 (90, 329) | 291,676 | 660 | 2.26 (2.10, 2.44) | 220 (103, 357) |
| Unknown | 1794953 | 1234 | 0.69 (0.65, 0.73) | 205 (90, 358) | 2409469 | 2806 | 1.16 (1.12, 1.21) | 233 (117, 370) |
| Rate of HgA1c Change in 1 Year |  |  |  |  |  |  |  |  |
| < -0.0008 | 126922 | 146 | 1.15 (0.97, 1.35) | 184 (88, 343) | 322572 | 519 | 1.61 (1.47, 1.75) | 227 (115, 371) |
| [-0.0008, 0.0008] | 280155 | 181 | 0.64 (0.56, 0.75) | 224 (98, 371) | 554791 | 535 | 0.96 (0.89, 1.05) | 249 (129, 378) |
| > 0.0008 | 129736 | 231 | 1.78 (1.56, 2.02) | 200 (92, 343) | 327480 | 722 | 2.21 (2.05, 2.37) | 225.5 (105, 359) |
| Unknown | 1794953 | 1234 | 0.69 (0.65, 0.73) | 205 (90, 358) | 2409469 | 2806 | 1.16 (1.12, 1.21) | 233 (117, 370) |
| Rate of Weight Change in 1 Year |  |  |  |  |  |  |  |  |
| < -0.02 | 503012 | 683 | 1.35 (1.26, 1.46) | 160 (68, 322) | 869902 | 2049 | 2.36 (2.26, 2.46) | 185 (86, 313) |
| [-0.02, 0.02] | 1033671 | 703 | 0.68 (0.63, 0.73) | 238 (124, 387) | 1437632 | 1403 | 0.98 (0.93, 1.03) | 286 (156, 409) |
| > 0.02 | 393142 | 209 | 0.53 (0.46, 0.61) | 238 (114, 358) | 750864 | 621 | 0.83 (0.76, 0.89) | 281 (145, 408) |
| Unknown | 401941 | 197 | 0.49 (0.43, 0.56) | 202 (90, 364) | 556817 | 509 | 0.91 (0.84, 0.996) | 244 (133, 387) |
| Abdominal Pain in Prior 6 mos<sup>b</sup> |  |  |  |  |  |  |  |  |
| Yes | 168579 | 322 | 1.91 (1.71, 2.13) | 116 (54, 256) | 139459 | 606 | 4.35 (4.01, 4.70) | 136 (67, 252) |
| No | 134525 | 139 | 1.03 (0.87, 1.22) | 199 (110, 349) | 3474756 | 3976 | 1.14 (1.11, 1.18) | 250.5 (128, 382.5) |
| Unknown | 2028662 | 1331 | 0.66 (0.62, 0.69) | 230 (110, 377) |  |  |  |  |

Abbreviations: ALT, alanine transaminase; CI, confidence interval; F/U, follow-up; HgA1c, hemoglobin A1c; IQR, interquartile range; PDAC, pancreatic ductal adenocarcinoma; PY, person-years.
<sup>a</sup> Not estimated for KPSC dataset due to the small number of events (39) in this group.
<sup>b</sup> For VA data, "Unknown" was interpreted as "No" due to the inability to distinguish between the two.

### Model Training, Validation and Testing

The number and size of the KPSC training, KPSC validation and VA testing datasets are shown in eTable 6.

For the main cohort, the preselection process identified 29 potential predictors (eTable 4). Of the 50 training samples, the three winning models containing age, abdominal pain, weight change and two biomarkers (alanine transaminase (ALT) change/HbA1c, rate of ALT change/HbA1c, and rate of ALT change/rate of HbA1c change) appeared most often (Table 3). Internal validation based on KPSC validation datasets revealed comparable results among the top three models (c-index: mean 0.77 for all three models (M1-M3) and SD 0.01-0.02; calibration test: p-value 0.2-0.4 and SD 0.2-0.3). When M1 was directly applied to VA testing datasets, the mean c-index was 0.69 (SD 0.003) (data not shown); however, after the algorithm was recalibrated based on VA’s datasets, the mean c-index based on 10 testing datasets was 0.71 (SD 0.002) (Table 3).

**Table 3.**
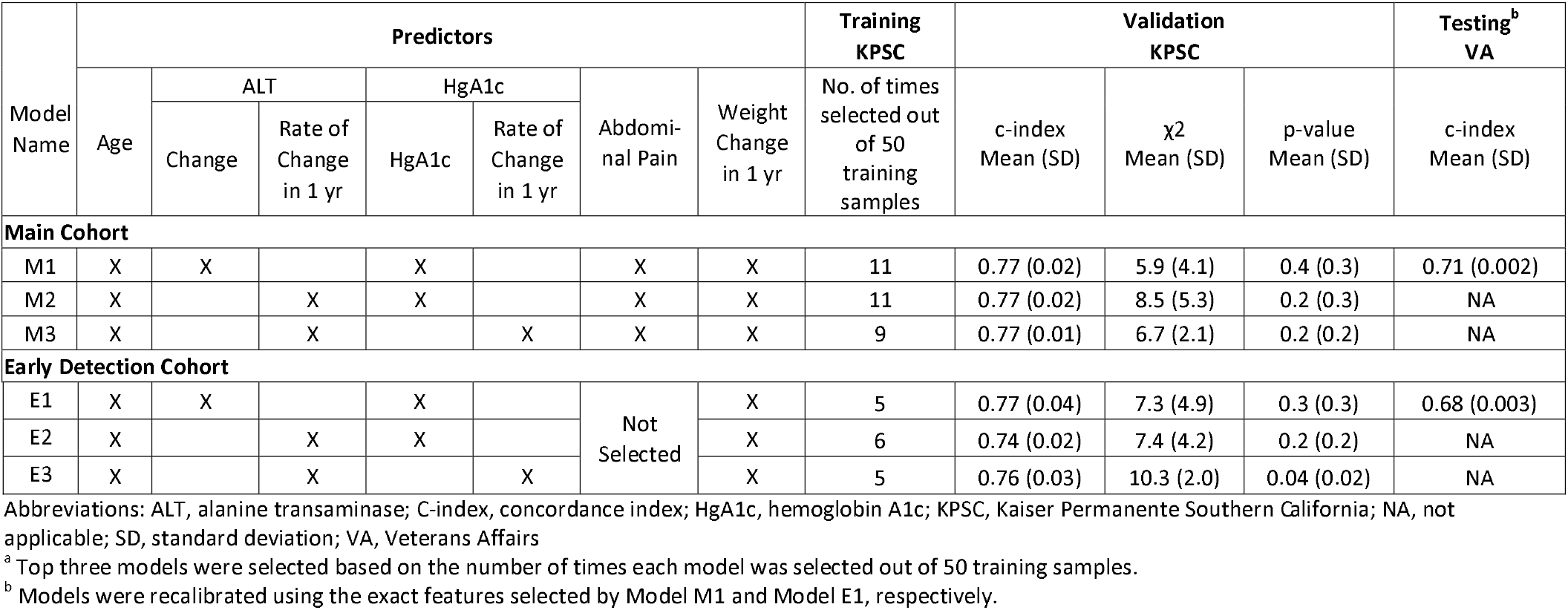
Predictors selected in top 3 models^a^ developed based on the main cohort and restricted cohort, and model performance during internal validation and external testing.

For the early detection cohort, the preselection process identified 32 potential predictors (eTable 4). The three best models (E1-E3, c-index: 0.74-0.77) contained the same features as those selected by M1-M3 except that abdominal pain was not chosen (Table 3). The calibration test for model E3 was significant (p=0.04), indicating a lack of model fit. The recalibrated E1 model based on the VA testing datasets achieved a mean c-index of 0.68 (SD 0.003) (Table 3).

The hyperparameters used and the features selected by ≥5 out of 50 models for each cohort can be found in eTables 7 and 8, respectively.

Figure 1 displays the calibration plots for all six models (M1-M3, E1-E3). It appears that all fit well for the four out of five lower risk groups (i.e., risk<95th percentile). However, for the highest risk group (risk ≥95th percentile), M1-M3 properly estimated the risks at KPSC, while E1-E3 slightly overestimated the risks for KPSC patients, and M1, E1 slightly underestimated the risks for VA patients.

**Figure 1.**
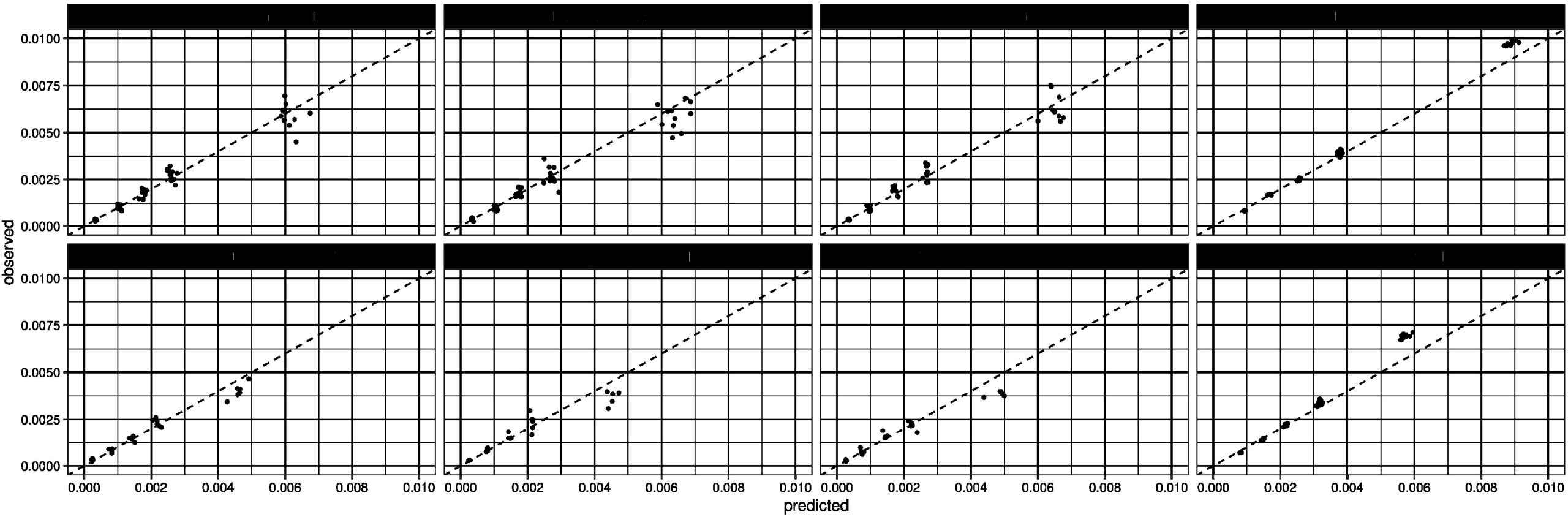
Calibration plots of winning models. x-axis: predicted; y-axis: observed. The five clusters represent the five risk groups defined by the ranges of predicted risks: <50th, 50–74th, 75–89th, 90–94th, 95–100th percentiles. Within each cluster, there are multiple dots representing the pairs of predicted and observed risks, calculated based on the corresponding validation datasets.

Sensitivity, specificity, PPV and fold increase in risk were comparable among the three winning models for both main cohort (M1-M3) and early detection cohort (E1-E3) (Table 4). The top 2.5% of the testing sample based on M1 experienced 1% risk of PDAC over 18 months, which was 7- to 8-fold higher than the baseline risk of PDAC in the KPSC cohort. Twenty % of the total PDAC cases occurring within 18 months were identified in this top 2.5% model-predicted high-risk group. Patients within the top 20% predicted risk of PDAC experienced 0.6% risk of PDAC over 18 months. This identified more than 50% of PDAC occurring within 18 months with a specificity of 80%. While sensitivities and fold increases in PDAC incidence rate were lower in the VA population, PPVs were higher for VA validation data compared to those of KPSC (Table 4).

**Table 4.** Percent of patients^a^ whose risk was among the top 20%, 15%, 10%, 5%, and 2.5%, sensitivity, specificity, positive predictive value (PPV), and risk fold increase for each of the winning models based on KPSC validation datasets and one of the winning models based on VA validation dataset.

|  | KPSC |  |  |  |  |  |  |  |  |  |  |  |  |  |  | VA |  |  |  |  |
| --- | --- | --- | --- | --- | --- | --- | --- | --- | --- | --- | --- | --- | --- | --- | --- | --- | --- | --- | --- | --- |
|  | High-Risk Patients |  |  |  |  | High-Risk Patients |  |  |  |  | High-Risk Patients |  |  |  |  | High-Risk Patients |  |  |  |  |
|  | Top 20% | Top 15% | Top 10% | Top 20% | Top 2.5% | Top 20% | Top 15% | Top 10% | Top 5% | Top 2.5% | Top 20% | Top 15% | Top 10% | Top 5% | Top 2.5% | Top 20% | Top 15% | Top 10% | Top 5% | Top 2.5% |
| **Main Cohort** | **Model M1** |  |  |  |  | **Model M2** |  |  |  |  | **Model M3** |  |  |  |  | **Model M1** |  |  |  |  |
| N<sup>b</sup> | 50952 | 37454 | 24959 | 424761 | 6249 | 49746 | 37512 | 24949 | 12572 | 6244 | 49934 | 37545 | 24850 | 12468 | 6238 | 424761 | 318408 | 212392 | 106118 | 53528 |
| Sensitivity (%) | 56.6 | 48.8 | 39.0 | 51.7 | 19.5 | 55.5 | 47.5 | 38.8 | 27.1 | 18.5 | 56.6 | 48.9 | 40.0 | 28.0 | 19.5 | 51.7 | 45.5 | 37.7 | 27.2 | 19.4 |
| Specificity (%) | 79.6 | 85.1 | 90.0 | 80.2 | 97.5 | 80.1 | 85.0 | 90.1 | 95.0 | 97.5 | 80.1 | 85.0 | 90.1 | 95.0 | 97.5 | 80.2 | 85.1 | 90.1 | 95.1 | 97.5 |
| PPV (%) | 0.4 | 0.5 | 0.6 | 0.6 | 1.1 | 0.4 | 0.4 | 0.5 | 0.8 | 1.0 | 0.4 | 0.5 | 0.6 | 0.8 | 1.1 | 0.6 | 0.7 | 0.8 | 1.2 | 1.7 |
| Fold Increase in Risk<sup>c</sup> | 2.8 | 3.3 | 3.9 | 2.6 | 7.9 | 2.8 | 3.2 | 3.9 | 5.4 | 7.4 | 2.8 | 3.3 | 4.0 | 5.6 | 7.9 | 2.6 | 3.0 | 3.7 | 5.4 | 7.7 |
| **Early Detection Cohort** | **Model E1** |  |  |  |  | **Model E2** |  |  |  |  | **Model E3** |  |  |  |  | **Model E1** |  |  |  |  |
| N<sup>b</sup> | 49944 | 37462 | 24974 | 424521 | 6244 | 49966 | 37475 | 24984 | 12492 | 6246 | 49917 | 37439 | 24960 | 12480 | 6240 | 424521 | 318131 | 212172 | 106087 | 53047 |
| Sensitivity (%) | 55.1 | 44.1 | 35.1 | 49.6 | 14.5 | 50.7 | 42.2 | 33.2 | 20.3 | 12.6 | 52.3 | 44.2 | 33.6 | 22.3 | 13.1 | 49.6 | 42.8 | 34.5 | 23.6 | 15.5 |
| Specificity (%) | 80.0 | 85.0 | 90.0 | 80.1 | 97.5 | 80.9 | 80.0 | 85.0 | 90.0 | 95.0 | 80.0 | 85.0 | 90.0 | 95.0 | 97.5 | 80.1 | 85.1 | 89.8 | 95.0 | 97.5 |
| PPV (%) | 0.3 | 0.3 | 0.4 | 0.4 | 0.6 | 0.3 | 0.3 | 0.4 | 0.4 | 0.5 | 0.3 | 0.3 | 0.4 | 0.5 | 0.6 | 0.4 | 0.6 | 0.6 | 0.8 | 1.1 |
| Fold Increase in Risk<sup>c</sup> | 2.6 | 2.9 | 3.5 | 2.5 | 5.8 | 2.5 | 2.8 | 3.3 | 4.1 | 5.0 | 2.6 | 2.9 | 3.4 | 4.5 | 5.2 | 2.5 | 2.9 | 3.5 | 4.7 | 6.2 |

Abbreviations: KPSC, Kaiser Permanente Southern California; PPV, positive predictive value; VA, Veterans Affairs.
<sup>a</sup> Estimated in patients with complete 18 months follow up or those who developed PDAC in 18 months.
<sup>b</sup> Number of eligible patients whose risk was above each risk threshold.
<sup>c</sup> Compared with the incidence rate in the entire cohort.
Model M1: Main cohort; age, weight change, abdominal pain, ALT change, HgA1c; estimated based on 11 validation samples for KPSC and 10 imputed testing datasets for VA.
Model M2: Main cohort; age, weight change, abdominal pain, ALT change rate, HgA1c; estimated based on 11 validation samples.
Model M3: Main cohort; age, weight change, abdominal pain, ALT change rate, HgA1c change rate; estimated based on 9 validation samples.
Model E1: Early detection cohort; age, weight change, ALT change, HgA1c; estimated based on 5 validation samples for KPSC and 10 imputed testing datasets for VA.
Model E2: Early detection cohort; age, weight change, ALT change rate, HgA1c, estimated based on 6 validation samples.
Model E3: Early detection cohort; age, weight change, ALT change rate, HgA1c change rate, estimated based on 5 validation samples.

Compared to the models developed based on the main cohort (M1-M3), the models developed based on early detection cohort (E1-E3) had slightly compromised sensitivity, PPV and fold increase in risk.

### Model application/implementation

A total of 606 patients were identified by the model with predicted risk of ≥1%. Nine (1.5%) patients had abnormalities identified in the region of the pancreas with the following diagnoses/stages: PDAC stages 1b, II, III, IV; pancreatic lymphoma; pancreatic neuroendocrine tumor; ampullary carcinoma; cholangiocarcinoma (n=2).

To facilitate external application of the RSF-based prediction model (M1), we have developed a publicly available web-based tool (https://pcriskdev.kp-scalresearch.org/). A hypothetical 70-year-old male patient with hemoglobin A1c value of 7.5%, weight loss of 4 lbs. and ALT increase of 4 IU/L in one year has an estimated 18-month risk of PDAC 0.30%.

For demonstration, decision rules based on one of the trees built for M1 is displayed in eFigures 2 and 3 for the left and the right side of the decision tree, respectively.

## DISCUSSION

We applied machine learning methods to EHR data to derive and validate clinical prediction models for sporadic pancreatic cancer across two large integrated healthcare systems. Despite inclusion of >500 potential features in the candidate pool, the machine learning models incorporated traditional parameters including age, glycated hemoglobin, alanine aminotransferase, weight, and abdominal pain. The final models were both parsimonious (with only 4-5 predictors) and reasonably accurate in both internal and external validation. In addition, the model proved well-calibrated when applied prospectively for real-time identification of not only pancreatic but biliary cancers.

While there has been some progress in studying approaches to early detection in high-risk patients based on either family history or genetic susceptibility^21^ as well as those with specific conditions such as late-onset diabetes,^6^ limited data exist on identification of patients at risk for sporadic pancreatic cancer. This study presents a novel approach to risk stratification at the population-level based on dynamic parameters contained within structured data from EHR.

Although parameters included in the model are well-established parameters for PDAC,^5,6,10^ their selection using an unbiased, comprehensive data-driven approach helped ensure inclusion of the most relevant combination of parameters. A recently developed model to predict risk of pancreatic cancer among patients with late or new onset diabetes at age 50 or later similarly identified increasing age, weight loss and change in blood glucose as key parameters for determining risk of pancreatic cancer in this patient population.^5^ The current model extends the concept of model-based risk prediction to a much broader population while maintaining reasonably high levels of discriminative accuracy for prediction of pancreatic cancer.

External validation is key to assessing model performance. In a review of 127 prediction models, Siontis et. al found 32 (25%) had at least one external validation.^22^ AUC estimates significantly decreased during external validation vs. the derivation study with a median AUC reduction 0.05.^22^ In the current study, the c-index declined by 0.08 (or 10%) when model M1 was directly transported and about 0.06 (or 8%) and 0.09 (or 12%) after models M1 and E1 were recalibrated, respectively. Although the comparison should be interpreted cautiously, the larger reduction observed in the current study could be attributable to multiple factors. First, a higher frequency of PDAC cases in the VA cohort were pancreatic cancer deaths identified through mortality records compared to the KPSC cohort. Second, given the differences in age and sex between KPSC and VA populations, a higher incidence rate of PDAC in the VA dataset compared to that of KP’s was observed as expected. This could have impacted model accuracy especially for the models without recalibration.

Strengths of the current study included a comprehensive, data-driven approach to model development, use of structured data elements and external validation in a separate healthcare system with distinct patient population. The present study also extended model development to assess feasibility of implementing the model through application in a prospective pilot study that demonstrated ability to identify pancreaticobiliary cancers in real-time.

The present study also had several limitations. First, several parameters identified in the prediction models (abdominal pain, abnormal ALT) are often associated with advanced stage pancreatic cancer. However, several early-stage pancreatic cancer cases were detected by the algorithm in the prospective feasibility study. Nevertheless, an ongoing concern relates to the timing with respect to clinical diagnosis. The 30-day cancer-free period used in the present model is likely insufficient to provide a reasonable window of opportunity for intervention to impact the disease course. To address this concern, we also developed an early detection model that restricted the study population to patients with ≥90 days cancer-free follow-up from the index date. Further testing of this restricted model for early detection of pancreatic cancer is the subject of an ongoing single-arm prospective interventional study (NCT04883450). Despite the reasonably high performance in terms of discriminative ability, the absolute risk in the highest risk category (top 2.5%) approached 1% over 18-months. This level of risk is likely below the threshold for cost-effective screening based on currently available testing.^23^ Second, of the 1479 and 4,582 events identified in the KPSC and VA cohorts, respectively, 300 and 2,564 events were captured by data sources other than Cancer Registry. An evaluation based on the KPSC Cancer Registry of the same time window showed that about 90% of pancreatic cancer cases were PDAC. Third, to estimate sensitivity, specificity, PPV and fold of risk increase, we relied on a subset of patients (∼70% and ∼80% of the total patients in the KPSC and VA cohorts, respectively) with complete follow up unless they died of pancreatic cancer. This restriction over-estimated the risk of PDAC, because the patients who were excluded from this analysis were at-risk for some periods of time. Finally, several cancers identified during prospective pilot testing of the algorithm were biliary in origin given similarities in anatomic location and clinical presentation.

In conclusion, we developed a parsimonious clinical risk prediction model for sporadic pancreatic cancer in a large, diverse integrated health system and subsequently applied the model in a separate health system. We also evaluated the model prospectively for real-time application. The model identified five key factors in determining risk of pancreatic cancer. Findings from the present study provide a potential framework for a systematic approach to targeted screening for pancreatic cancer based on automated analysis of data in EHR.

## Supporting information

eFigure 1

eFigure 2

eFigure 3

## Data Availability

Anonymized data that support the findings of this study may be made available from the investigative team in the following conditions: 1) agreement to collaborate with the study team on all publications, 2) provision of external funding for administrative and investigator time necessary for this collaboration, 3) demonstration that the external investigative team is qualified and has documented evidence of training for human subjects protections, and 4) agreement to abide by the terms outlined in data use agreements between institutions.

## Abbreviations

ALT: Alanine transaminase
AUC: area under the curve
CI: confidence interval
DoD: Department of Defense
EHR: electronic health record
GND: Greenwood-Nam-D’Agostino
HbA1C: glycated hemoglobin
ICD-9-CM: Ninth Revision of International Classification of Diseases, Clinical Modification
ICD-10-CM: Tenth Revision of International Classification of Diseases, Clinical Modification
IR: Incidence rate
KPSC: Kaiser Permanente Southern California
NDI: National Death Index
NOD: new onset diabetes
PC: pancreatic cancer
PDAC: pancreatic ductal adenocarcinoma
PPV: positive predictive value
RSF: Random Survival Forest
SEER: Surveillance, Epidemiology, and End Results
VA: Veterans Affairs

## Figure Legends

**eFigure 1** – Consort Diagram (KPSC and VA main cohorts)

**eFigure 2** – One of the decision trees for KPSC M1 (left side)

**eFigure 3** – One of the decision trees for KPSC M1 (right side)

## Notes

**Author Conflict of Interest / Study Support**

**Financial support:** Research reported in this publication was supported by the National Cancer Institute of the National Institutes of Health under Award Number R01CA230442. The content is solely the responsibility of the authors and does not necessarily represent the official views of the National Institutes of Health.

**Potential competing interests:** The authors declare they have no conflict of interest for this study.

### Competing Interest Statement

The authors have declared no competing interest.

### Funding Statement

Research reported in this publication was supported by the National Cancer Institute of the National Institutes of Health under Award Number R01CA230442. The content is solely the responsibility of the authors and does not necessarily represent the official views of the National Institutes of Health.

### Author Declarations

Kaiser Permanente Southern California's Institutional Review Board gave ethical approval for this work.

