## Supplementary material for "Prediction Model for Detection of Sporadic Pancreatic Cancer (PRO-TECT) in a Population-Based Cohort Using Machine Learning and Further Validation in a Prospective Study": Online Supplements

**Online Supplements: Online Tables eTable 1- eTable 7**

**Table of Contents**

**eTable 1. ICD‑O‑3 Histology Codes Used to Identify Pancreatic Ductal Adenocarcinoma (PDAC).**

**eTable 2. Potential features included in the study.**

**eTable 3: Definitions of weight/laboratory value changes and symptom related features.**

**eTable 4. Feature selection process and final preselected potential features for the main and early detection cohorts.**

**eTable 5. Distribution of American Joint Committee on Cancer (AJCC) stage in patients with PDAC.**

**eTable 6. Number and size of the KPSC training, KPSC validation and VA testing datasets.**

**eTable 7. Hyperparameter setup for both feature pre-selection and model development for the main and early detectiond cohorts.**

**eTable 8. Predictors selected by at least 5 out of the 50 models based on the 50 training samples.**

**eTable 1. ICD‑O‑3 Histology Codes Used to Identify Pancreatic Ductal Adenocarcinoma (PDAC).**

8000

8001

8010

8020

8021

8022

8140

8141

8143

8210

8211

8230

8255

8500

8501

8503

8504

8507

8508

8521

8552

8560

8570

8571

8572

8573

8574

8575

8576

Please refer to the Web site for information on the histology codes.

[https://www.naaccr.org/wp-content/uploads/2018/01/](https://www.naaccr.org/wp-content/uploads/2018/01/Updated-Jan-10-2018-ICD-O-3-Guidelines-v2.pdf) [Updated-Jan-10-2018-ICD-O-3-Guidelines-v2.pdf](https://www.naaccr.org/wp-content/uploads/2018/01/Updated-Jan-10-2018-ICD-O-3-Guidelines-v2.pdf).

**eTable 2. Potential features included in the study.**

| **Feature Category** | **Variable** |
| --- | --- |
| Demographics | age at index date |
| Demographics | diagnosis of alcohol abuse in year prior to index date |
| Demographics | diagnosis of alcohol abuse ever |
| Demographics | maximum reported alcohol use per week ever |
| Demographics | BMI closest and prior to index date |
| Demographics | maximum reported BMI ever |
| Demographics | personal history of any cancer prior to index |
| Demographics | reported minutes of exercise per week, closest and prior to index |
| Demographics | reported family history (noted at any time prior to index date) of any cancer |
| Demographics | reported family history (noted at any time prior to index date) of breast cancer |
| Demographics | reported family history (noted at any time prior to index date) of colon cancer |
| Demographics | reported family history (noted at any time prior to index date) of ovarian cancer |
| Demographics | reported family history (noted at any time prior to index date) of prostate cancer |
| Demographics | reported family history (noted at any time prior to index date) of pancreatic cancer |
| Demographics | gender |
| Demographics | maximum reported height ever |
| Demographics | race/ethnicity - all categories |
| Demographics | response or "quit" or "ever" to tobacco use ever prior to index date |
| Demographics | maximum years or reported tobacco use prior to index date |
| Demographics | weight closest and prior to index date |
| Demographics | maximum reported weight ever |
| Demographics | weight change per day, calculated from the two farthest values within the year prior to index date |
| Demographics | test indicating Jewish ancestry ever prior to index date |
| Demographics | ever indication of Jewish religion (collected from inpatient) |
| Demographics | reported census tract at index date |
| Demographics | geocoded latitude of home address at index date |
| Demographics | geocoded longitude of home address at index date |
| Diagnoses | acute pancreatitis, between 0-6 months prior to index date |
| Diagnoses | acute pancreatitis, between 7-12 months prior to index date |
| Diagnoses | acute pancreatitis, between 1-2 years prior to index date |
| Diagnoses | acute pancreatitis, more than 2 years prior to index date |
| Diagnoses | anxiety, between 0-6 months prior to index date |
| Diagnoses | anxiety, between 7-12 months prior to index date |
| Diagnoses | anxiety, between 1-2 years prior to index date |
| Diagnoses | anxiety, more than 2 years prior to index date |
| Diagnoses | benign pancreatic disorders, between 0-6 months prior to index date |
| Diagnoses | benign pancreatic disorders, between 7-12 months prior to index date |
| Diagnoses | benign pancreatic disorders, between 1-2 years prior to index date |
| Diagnoses | benign pancreatic disorders, more than 2 years prior to index date |
| Diagnoses | biliary tract disease, between 0-6 months prior to index date |
| Diagnoses | biliary tract disease, between 7-12 months prior to index date |
| Diagnoses | biliary tract disease, between 1-2 years prior to index date |
| Diagnoses | biliary tract disease, more than 2 years prior to index date |
| Diagnoses | cardiovascular disease, between 0-6 months prior to index date |
| Diagnoses | cardiovascular disease, between 7-12 months prior to index date |
| Diagnoses | cardiovascular disease, between 1-2 years prior to index date |
| Diagnoses | cardiovascular disease, more than 2 years prior to index date |
| Diagnoses | chronic pancreatitis, between 0-6 months prior to index date |
| Diagnoses | chronic pancreatitis, between 7-12 months prior to index date |
| Diagnoses | chronic pancreatitis, between 1-2 years prior to index date |
| Diagnoses | chronic pancreatitis, more than 2 years prior to index date |
| Diagnoses | colorectal polyps, between 0-6 months prior to index date |
| Diagnoses | colorectal polyps, between 7-12 months prior to index date |
| Diagnoses | colorectal polyps, between 1-2 years prior to index date |
| Diagnoses | colorectal polyps, more than 2 years prior to index date |
| Diagnoses | chronic pulmonary disease, between 0-6 months prior to index date |
| Diagnoses | chronic pulmonary disease, between 7-12 months prior to index date |
| Diagnoses | chronic pulmonary disease, between 1-2 years prior to index date |
| Diagnoses | chronic pulmonary disease, more than 2 years prior to index date |
| Diagnoses | Crohn's disease, between 0-6 months prior to index date |
| Diagnoses | Crohn's disease, between 7-12 months prior to index date |
| Diagnoses | Crohn's disease, between 1-2 years prior to index date |
| Diagnoses | Crohn's disease, more than 2 years prior to index date |
| Diagnoses | cystic fibrosis, between 0-6 months prior to index date |
| Diagnoses | cystic fibrosis, between 7-12 months prior to index date |
| Diagnoses | cystic fibrosis, between 1-2 years prior to index date |
| Diagnoses | cystic fibrosis, more than 2 years prior to index date |
| Diagnoses | depression, between 0-6 months prior to index date |
| Diagnoses | depression, between 7-12 months prior to index date |
| Diagnoses | depression, between 1-2 years prior to index date |
| Diagnoses | depression, more than 2 years prior to index date |
| Diagnoses | dermatologic conditions, between 0-6 months prior to index date |
| Diagnoses | dermatologic conditions, between 7-12 months prior to index date |
| Diagnoses | dermatologic conditions, between 1-2 years prior to index date |
| Diagnoses | dermatologic conditions, more than 2 years prior to index date |
| Diagnoses | diabetes mellitus, between 0-6 months prior to index date |
| Diagnoses | diabetes mellitus, between 7-12 months prior to index date |
| Diagnoses | diabetes mellitus, between 1-2 years prior to index date |
| Diagnoses | diabetes mellitus, more than 2 years prior to index date |
| Diagnoses | diabetes complications of ketoacidosis, coma, or hyperosmolarity, between 0-6 months prior to index date |
| Diagnoses | diabetes complications of ketoacidosis, coma, or hyperosmolarity, between 7-12 months prior to index date |
| Diagnoses | diabetes complications of ketoacidosis, coma, or hyperosmolarity, between 1-2 years prior to index date |
| Diagnoses | diabetes complications of ketoacidosis, coma, or hyperosmolarity, more than 2 years prior to index date |
| Diagnoses | diabetes complications, neurological or ophthalmic, between 0-6 months prior to index date |
| Diagnoses | diabetes complications, neurological or ophthalmic, between 7-12 months prior to index date |
| Diagnoses | diabetes complications, neurological or ophthalmic, between 1-2 years prior to index date |
| Diagnoses | diabetes complications, neurological or ophthalmic, more than 2 years prior to index date |
| Diagnoses | diabetes complications, circulatory or kidney, between 0-6 months prior to index date |
| Diagnoses | diabetes complications, circulatory or kidney, between 7-12 months prior to index date |
| Diagnoses | diabetes complications, circulatory or kidney, between 1-2 years prior to index date |
| Diagnoses | diabetes complications, circulatory or kidney, more than 2 years prior to index date |
| Diagnoses | other diabetes complications, between 0-6 months prior to index date |
| Diagnoses | other diabetes complications, between 7-12 months prior to index date |
| Diagnoses | other diabetes complications, between 1-2 years prior to index date |
| Diagnoses | other diabetes complications, circulatory or kidney, more than 2 years prior to index date |
| Diagnoses | type of diabetes mellitus, between 0-6 months prior to index date |
| Diagnoses | type of diabetes mellitus, between 7-12 months prior to index date |
| Diagnoses | type of diabetes mellitus, between 1-2 years prior to index date |
| Diagnoses | type of diabetes mellitus, more than 2 years prior to index date |
| Diagnoses | dyspepsia, between 0-6 months prior to index date |
| Diagnoses | dyspepsia, between 7-12 months prior to index date |
| Diagnoses | dyspepsia, between 1-2 years prior to index date |
| Diagnoses | dyspepsia, more than 2 years prior to index date |
| Diagnoses | eosinophilia, between 0-6 months prior to index date |
| Diagnoses | eosinophilia, between 7-12 months prior to index date |
| Diagnoses | eosinophilia, between 1-2 years prior to index date |
| Diagnoses | eosinophilia, more than 2 years prior to index date |
| Diagnoses | eosinophilis esophagitis, between 0-6 months prior to index date |
| Diagnoses | eosinophilis esophagitis, between 7-12 months prior to index date |
| Diagnoses | eosinophilis esophagitis, between 1-2 years prior to index date |
| Diagnoses | eosinophilis esophagitis, more than 2 years prior to index date |
| Diagnoses | gallstone cholangitis, between 0-6 months prior to index date |
| Diagnoses | gallstone cholangitis, between 7-12 months prior to index date |
| Diagnoses | gallstone cholangitis, between 1-2 years prior to index date |
| Diagnoses | gallstone cholangitis, more than 2 years prior to index date |
| Diagnoses | gastritis, between 0-6 months prior to index date |
| Diagnoses | gastritis, between 7-12 months prior to index date |
| Diagnoses | gastritis, between 1-2 years prior to index date |
| Diagnoses | gastritis, more than 2 years prior to index date |
| Diagnoses | granulomatosis with polyangiitis, between 0-6 months prior to index date |
| Diagnoses | granulomatosis with polyangiitis, between 7-12 months prior to index date |
| Diagnoses | granulomatosis with polyangiitis, between 1-2 years prior to index date |
| Diagnoses | granulomatosis with polyangiitis, more than 2 years prior to index date |
| Diagnoses | hepatobiliary disease, between 0-6 months prior to index date |
| Diagnoses | hepatobiliary disease, between 7-12 months prior to index date |
| Diagnoses | hepatobiliary disease, between 1-2 years prior to index date |
| Diagnoses | hepatobiliary disease, more than 2 years prior to index date |
| Diagnoses | hereditary cancer syndrome, between 0-6 months prior to index date |
| Diagnoses | hereditary cancer syndrome, between 7-12 months prior to index date |
| Diagnoses | hereditary cancer syndrome, between 1-2 years prior to index date |
| Diagnoses | hereditary cancer syndrome, more than 2 years prior to index date |
| Diagnoses | HIV, between 0-6 months prior to index date |
| Diagnoses | HIV, between 7-12 months prior to index date |
| Diagnoses | HIV, between 1-2 years prior to index date |
| Diagnoses | HIV, more than 2 years prior to index date |
| Diagnoses | hypertension, between 0-6 months prior to index date |
| Diagnoses | hypertension, between 7-12 months prior to index date |
| Diagnoses | hypertension, between 1-2 years prior to index date |
| Diagnoses | hypertension, more than 2 years prior to index date |
| Diagnoses | immunodeficiency disorders, between 0-6 months prior to index date |
| Diagnoses | immunodeficiency disorders, between 7-12 months prior to index date |
| Diagnoses | immunodeficiency disorders, between 1-2 years prior to index date |
| Diagnoses | immunodeficiency disorders, more than 2 years prior to index date |
| Diagnoses | lupus erythematosus, between 0-6 months prior to index date |
| Diagnoses | lupus erythematosus, between 7-12 months prior to index date |
| Diagnoses | lupus erythematosus, between 1-2 years prior to index date |
| Diagnoses | lupus erythematosus, more than 2 years prior to index date |
| Diagnoses | mast cell disease, between 0-6 months prior to index date |
| Diagnoses | mast cell disease, between 7-12 months prior to index date |
| Diagnoses | mast cell disease, between 1-2 years prior to index date |
| Diagnoses | mast cell disease, more than 2 years prior to index date |
| Diagnoses | non-alcoholic fatty liver disease, between 0-6 months prior to index date |
| Diagnoses | non-alcoholic fatty liver disease, between 7-12 months prior to index date |
| Diagnoses | non-alcoholic fatty liver disease, between 1-2 years prior to index date |
| Diagnoses | non-alcoholic fatty liver disease, more than 2 years prior to index date |
| Diagnoses | pulmonary circulation disorder, between 0-6 months prior to index date |
| Diagnoses | pulmonary circulation disorder, between 7-12 months prior to index date |
| Diagnoses | pulmonary circulation disorder, between 1-2 years prior to index date |
| Diagnoses | pulmonary circulation disorder, more than 2 years prior to index date |
| Diagnoses | peptic ulcer, between 0-6 months prior to index date |
| Diagnoses | peptic ulcer, between 7-12 months prior to index date |
| Diagnoses | peptic ulcer, between 1-2 years prior to index date |
| Diagnoses | peptic ulcer, more than 2 years prior to index date |
| Diagnoses | prediabetes, between 0-6 months prior to index date |
| Diagnoses | prediabetes, between 7-12 months prior to index date |
| Diagnoses | prediabetes, between 1-2 years prior to index date |
| Diagnoses | prediabetes, more than 2 years prior to index date |
| Diagnoses | relapsing polychondritis, between 0-6 months prior to index date |
| Diagnoses | relapsing polychondritis, between 7-12 months prior to index date |
| Diagnoses | relapsing polychondritis, between 1-2 years prior to index date |
| Diagnoses | relapsing polychondritis, more than 2 years prior to index date |
| Diagnoses | renal disease, between 0-6 months prior to index date |
| Diagnoses | renal disease, between 7-12 months prior to index date |
| Diagnoses | renal disease, between 1-2 years prior to index date |
| Diagnoses | renal disease, more than 2 years prior to index date |
| Diagnoses | rheumatoid arthritis, between 0-6 months prior to index date |
| Diagnoses | rheumatoid arthritis, between 7-12 months prior to index date |
| Diagnoses | rheumatoid arthritis, between 1-2 years prior to index date |
| Diagnoses | rheumatoid arthritis, more than 2 years prior to index date |
| Diagnoses | sarcoidosis, between 0-6 months prior to index date |
| Diagnoses | sarcoidosis, between 7-12 months prior to index date |
| Diagnoses | sarcoidosis, between 1-2 years prior to index date |
| Diagnoses | sarcoidosis, more than 2 years prior to index date |
| Diagnoses | Charlson comorbidity index within one year prior to index date |
| Lab | alpha-1-fetoprotein, rate of change in lab result between the most recent and second most recent lab |
| Lab | alpha-1-fetoprotein, most recent lab result |
| Lab | alpha-1-fetoprotein, second most recent lab result |
| Lab | albumin, rate of change in lab result between the most recent and second most recent lab |
| Lab | albumin, most recent lab result |
| Lab | albumin, second most recent lab result |
| Lab | alkaline phosphatase, rate of change in lab result between the most recent and second most recent lab |
| Lab | alkaline phosphatase, most recent lab result |
| Lab | alkaline phosphatase, second most recent lab result |
| Lab | alanine aminotransferase, rate of change in lab result between the most recent and second most recent lab |
| Lab | alanine aminotransferase, most recent lab result |
| Lab | alanine aminotransferase, second most recent lab result |
| Lab | amylase, rate of change in lab result between the most recent and second most recent lab |
| Lab | amylase, most recent lab result |
| Lab | amylase, second most recent lab result |
| Lab | antinuclear antibody (ANA) titer result, between 0-6 months prior to index date |
| Lab | antinuclear antibody (ANA) titer result, between 7-12 months prior to index date |
| Lab | antinuclear antibody (ANA) titer result, between 1-2 years prior to index date |
| Lab | antinuclear antibody (ANA) titer result, more than 2 years prior to index date |
| Lab | neutrophils in blood, rate of change in lab result between the most recent and second most recent lab |
| Lab | neutrophils in blood, most recent lab result |
| Lab | neutrophils in blood, second most recent lab result |
| Lab | aspartate aminotransferase, rate of change in lab result between the most recent and second most recent lab |
| Lab | aspartate aminotransferase, most recent lab result |
| Lab | aspartate aminotransferase, second most recent lab result |
| Lab | direct bilirubin, rate of change in lab result between the most recent and second most recent lab |
| Lab | direct bilirubin, most recent lab result |
| Lab | direct bilirubin, second most recent lab result |
| Lab | total bilirubin, rate of change in lab result between the most recent and second most recent lab |
| Lab | total bilirubin, most recent lab result |
| Lab | total bilirubin, second most recent lab result |
| Lab | blood type result, between 0-6 months prior to index date |
| Lab | blood type result, between 7-12 months prior to index date |
| Lab | blood type result, between 1-2 years prior to index date |
| Lab | blood type result, more than 2 years prior to index date |
| Lab | brain natriuretic peptide, rate of change in lab result between the most recent and second most recent lab |
| Lab | brain natriuretic peptide, most recent lab result |
| Lab | brain natriuretic peptide, second most recent lab result |
| Lab | urea nitrogen in blood/serum, rate of change in lab result between the most recent and second most recent lab |
| Lab | urea nitrogen in blood/serum, most recent lab result |
| Lab | urea nitrogen in blood/serum, second most recent lab result |
| Lab | cancer antigen 125, rate of change in lab result between the most recent and second most recent lab |
| Lab | cancer antigen 125, most recent lab result |
| Lab | cancer antigen 125, second most recent lab result |
| Lab | carbohydrate antigen 19-9, rate of change in lab result between the most recent and second most recent lab |
| Lab | carbohydrate antigen 19-9, most recent lab result |
| Lab | carbohydrate antigen 19-9, second most recent lab result |
| Lab | calcium, rate of change in lab result between the most recent and second most recent lab |
| Lab | calcium, most recent lab result |
| Lab | calcium, second most recent lab result |
| Lab | carcinoembryonic antigen, rate of change in lab result between the most recent and second most recent lab |
| Lab | carcinoembryonic antigen, most recent lab result |
| Lab | carcinoembryonic antigen, second most recent lab result |
| Lab | celiac transglutaminase result, between 0-6 months prior to index date |
| Lab | celiac transglutaminase result, between 7-12 months prior to index date |
| Lab | celiac transglutaminase result, between 1-2 years prior to index date |
| Lab | celiac transglutaminase result, more than 2 years prior to index date |
| Lab | chloride, rate of change in lab result between the most recent and second most recent lab |
| Lab | chloride, most recent lab result |
| Lab | chloride, second most recent lab result |
| Lab | creatine kinase, rate of change in lab result between the most recent and second most recent lab |
| Lab | creatine kinase, most recent lab result |
| Lab | creatine kinase, second most recent lab result |
| Lab | cytomegalovirus result, between 0-6 months prior to index date |
| Lab | cytomegalovirus result, between 7-12 months prior to index date |
| Lab | cytomegalovirus result, between 1-2 years prior to index date |
| Lab | cytomegalovirus result, more than 2 years prior to index date |
| Lab | serum creatinine, rate of change in lab result between the most recent and second most recent lab |
| Lab | serum creatinine, most recent lab result |
| Lab | serum creatinine, second most recent lab result |
| Lab | c-reactive protein, rate of change in lab result between the most recent and second most recent lab |
| Lab | c-reactive protein, most recent lab result |
| Lab | c-reactive protein, second most recent lab result |
| Lab | high-sensitivity c-reactive protein, rate of change in lab result between the most recent and second most recent lab |
| Lab | high-sensitivity c-reactive protein, most recent lab result |
| Lab | high-sensitivity c-reactive protein, second most recent lab result |
| Lab | D-dimer fibrin derivatives, fibrinogen equivalent units, rate of change in lab result between the most recent and second most recent lab |
| Lab | D-dimer fibrin derivatives, fibrinogen equivalent units, most recent lab result |
| Lab | D-dimer fibrin derivatives, fibrinogen equivalent units, second most recent lab result |
| Lab | D-dimer fibrin derivatives, not specified, rate of change in lab result between the most recent and second most recent lab |
| Lab | D-dimer fibrin derivatives, not specified, most recent lab result |
| Lab | D-dimer fibrin derivatives, not specified, second most recent lab result |
| Lab | D-dimer fibrin derivatives, qualitative, rate of change in lab result between the most recent and second most recent lab |
| Lab | D-dimer fibrin derivatives, qualitative, most recent lab result |
| Lab | D-dimer fibrin derivatives, qualitative, second most recent lab result |
| Lab | Epstein-Barr virus result, between 0-6 months prior to index date |
| Lab | Epstein-Barr virus result, between 7-12 months prior to index date |
| Lab | Epstein-Barr virus result, between 1-2 years prior to index date |
| Lab | Epstein-Barr virus result, more than 2 years prior to index date |
| Lab | eosinophils (#/volume) in blood, rate of change in lab result between the most recent and second most recent lab |
| Lab | eosinophils (#/volume) in blood, most recent lab result |
| Lab | eosinophils (#/volume) in blood, second most recent lab result |
| Lab | eosinophils (#/volume) in blood, automated count, rate of change in lab result between the most recent and second most recent lab |
| Lab | eosinophils (#/volume) in blood, automated count, most recent lab result |
| Lab | eosinophils (#/volume) in blood, automated count, second most recent lab result |
| Lab | eosinophils (#/volume) in blood, manual count, rate of change in lab result between the most recent and second most recent lab |
| Lab | eosinophils (#/volume) in blood, manual count, most recent lab result |
| Lab | eosinophils (#/volume) in blood, manual count, second most recent lab result |
| Lab | eosinophils/100 leukocytes in blood, automated count, rate of change in lab result between the most recent and second most recent lab |
| Lab | eosinophils/100 leukocytes in blood, automated count, most recent lab result |
| Lab | eosinophils/100 leukocytes in blood, automated count, second most recent lab result |
| Lab | eosinophils/100 leukocytes in blood, manual count, rate of change in lab result between the most recent and second most recent lab |
| Lab | eosinophils/100 leukocytes in blood, manual count, most recent lab result |
| Lab | eosinophils/100 leukocytes in blood, manual count, second most recent lab result |
| Lab | erythrocyte sedimentation rate, rate of change in lab result between the most recent and second most recent lab |
| Lab | erythrocyte sedimentation rate, most recent lab result |
| Lab | erythrocyte sedimentation rate, second most recent lab result |
| Lab | ferritin, rate of change in lab result between the most recent and second most recent lab |
| Lab | ferritin, most recent lab result |
| Lab | ferritin, second most recent lab result |
| Lab | fibrinogen, rate of change in lab result between the most recent and second most recent lab |
| Lab | fibrinogen, most recent lab result |
| Lab | fibrinogen, second most recent lab result |
| Lab | fecal immunochemical test result, between 0-6 months prior to index date |
| Lab | fecal immunochemical test result, between 7-12 months prior to index date |
| Lab | fecal immunochemical test result, between 1-2 years prior to index date |
| Lab | fecal immunochemical test result, more than 2 years prior to index date |
| Lab | fecal occult blood result, between 0-6 months prior to index date |
| Lab | fecal occult blood result, between 7-12 months prior to index date |
| Lab | fecal occult blood result, between 1-2 years prior to index date |
| Lab | fecal occult blood result, more than 2 years prior to index date |
| Lab | folate, rate of change in lab result between the most recent and second most recent lab |
| Lab | folate, most recent lab result |
| Lab | folate, second most recent lab result |
| Lab | folate in red blood cells, rate of change in lab result between the most recent and second most recent lab |
| Lab | folate in red blood cells, most recent lab result |
| Lab | folate in red blood cells, second most recent lab result |
| Lab | gamma glutamyl transpeptidase or transferase in blood/serum, rate of change in lab result between the most recent and second most recent lab |
| Lab | gamma glutamyl transpeptidase or transferase in blood/serum, most recent lab result |
| Lab | gamma glutamyl transpeptidase or transferase in blood/serum, second most recent lab result |
| Lab | glucose, fasting serum/plasma, rate of change in lab result between the most recent and second most recent lab |
| Lab | glucose, fasting serum/plasma, most recent lab result |
| Lab | glucose, fasting serum/plasma, second most recent lab result |
| Lab | glucose, random, rate of change in lab result between the most recent and second most recent lab |
| Lab | glucose, random, most recent lab result |
| Lab | glucose, random, second most recent lab result |
| Lab | hematocrit in blood, rate of change in lab result between the most recent and second most recent lab |
| Lab | hematocrit in blood, most recent lab result |
| Lab | hematocrit in blood, second most recent lab result |
| Lab | high-density lipoprotein cholesterol, rate of change in lab result between the most recent and second most recent lab |
| Lab | high-density lipoprotein cholesterol, most recent lab result |
| Lab | high-density lipoprotein cholesterol, second most recent lab result |
| Lab | hepatitis B result, between 0-6 months prior to index date |
| Lab | hepatitis B result, between 7-12 months prior to index date |
| Lab | hepatitis B result, between 1-2 years prior to index date |
| Lab | hepatitis B result, more than 2 years prior to index date |
| Lab | hepatitis C result, between 0-6 months prior to index date |
| Lab | hepatitis C result, between 7-12 months prior to index date |
| Lab | hepatitis C result, between 1-2 years prior to index date |
| Lab | hepatitis C result, more than 2 years prior to index date |
| Lab | hemoglobin, rate of change in lab result between the most recent and second most recent lab |
| Lab | hemoglobin, most recent lab result |
| Lab | hemoglobin, second most recent lab result |
| Lab | hemoglobin A1c, rate of change in lab result between the most recent and second most recent lab |
| Lab | hemoglobin A1c, most recent lab result |
| Lab | hemoglobin A1c, second most recent lab result |
| Lab | human papillomavirus result, between 0-6 months prior to index date |
| Lab | human papillomavirus result, between 7-12 months prior to index date |
| Lab | human papillomavirus result, between 1-2 years prior to index date |
| Lab | human papillomavirus result, more than 2 years prior to index date |
| Lab | current (active) infection of Helicobacter pylori result, between 0-6 months prior to index date |
| Lab | current (active) infection of Helicobacter pylori result, between 7-12 months prior to index date |
| Lab | current (active) infection of Helicobacter pylori result, between 1-2 years prior to index date |
| Lab | current (active) infection of Helicobacter pylori result, more than 2 years prior to index date |
| Lab | historical infection of Helicobacter pylori result, between 0-6 months prior to index date |
| Lab | historical infection of Helicobacter pylori result, between 7-12 months prior to index date |
| Lab | historical infection of Helicobacter pylori result, between 1-2 years prior to index date |
| Lab | historical infection of Helicobacter pylori result, more than 2 years prior to index date |
| Lab | herpes simplex virus result, between 0-6 months prior to index date |
| Lab | herpes simplex virus result, between 7-12 months prior to index date |
| Lab | herpes simplex virus result, between 1-2 years prior to index date |
| Lab | herpes simplex virus result, more than 2 years prior to index date |
| Lab | immunoglobulin A, rate of change in lab result between the most recent and second most recent lab |
| Lab | immunoglobulin A, most recent lab result |
| Lab | immunoglobulin A, second most recent lab result |
| Lab | immunoglobulin G, rate of change in lab result between the most recent and second most recent lab |
| Lab | immunoglobulin G, most recent lab result |
| Lab | immunoglobulin G, second most recent lab result |
| Lab | immunoglobulin M, rate of change in lab result between the most recent and second most recent lab |
| Lab | immunoglobulin M, most recent lab result |
| Lab | immunoglobulin M, second most recent lab result |
| Lab | international normalization ratio, rate of change in lab result between the most recent and second most recent lab |
| Lab | international normalization ratio, most recent lab result |
| Lab | international normalization ratio, second most recent lab result |
| Lab | serum potassium, rate of change in lab result between the most recent and second most recent lab |
| Lab | serum potassium, most recent lab result |
| Lab | serum potassium, second most recent lab result |
| Lab | lactase dehydrogenase, rate of change in lab result between the most recent and second most recent lab |
| Lab | lactase dehydrogenase, most recent lab result |
| Lab | lactase dehydrogenase, second most recent lab result |
| Lab | low-density lipoprotein cholesterol, calculated, not specified, rate of change in lab result between the most recent and second most recent lab |
| Lab | low-density lipoprotein cholesterol, calculated, not specified, most recent lab result |
| Lab | low-density lipoprotein cholesterol, calculated, not specified, second most recent lab result |
| Lab | low-density lipoprotein cholesterol, direct measure, rate of change in lab result between the most recent and second most recent lab |
| Lab | low-density lipoprotein cholesterol, direct measure, most recent lab result |
| Lab | low-density lipoprotein cholesterol, direct measure, second most recent lab result |
| Lab | low-density lipoprotein cholesterol, not specified, rate of change in lab result between the most recent and second most recent lab |
| Lab | low-density lipoprotein cholesterol, not specified, most recent lab result |
| Lab | low-density lipoprotein cholesterol, not specified, second most recent lab result |
| Lab | lipase, rate of change in lab result between the most recent and second most recent lab |
| Lab | lipase, most recent lab result |
| Lab | lipase, second most recent lab result |
| Lab | lymphocytes (#/volume) in blood, rate of change in lab result between the most recent and second most recent lab |
| Lab | lymphocytes (#/volume) in blood, most recent lab result |
| Lab | lymphocytes (#/volume) in blood, second most recent lab result |
| Lab | lymphocytes (#/volume) in blood by automated count, rate of change in lab result between the most recent and second most recent lab |
| Lab | lymphocytes (#/volume) in blood by automated count, most recent lab result |
| Lab | lymphocytes (#/volume) in blood by automated count, second most recent lab result |
| Lab | lymphocytes (#/volume) in blood by manual count, rate of change in lab result between the most recent and second most recent lab |
| Lab | lymphocytes (#/volume) in blood by manual count, most recent lab result |
| Lab | lymphocytes (#/volume) in blood by manual count, second most recent lab result |
| Lab | lymphocytes/100 leukocytes in body fluid, rate of change in lab result between the most recent and second most recent lab |
| Lab | lymphocytes/100 leukocytes in body fluid, most recent lab result |
| Lab | lymphocytes/100 leukocytes in body fluid, second most recent lab result |
| Lab | lymphocytes/100 leukocytes in blood by automated count, rate of change in lab result between the most recent and second most recent lab |
| Lab | lymphocytes/100 leukocytes in blood by automated count, most recent lab result |
| Lab | lymphocytes/100 leukocytes in blood by automated count, second most recent lab result |
| Lab | lymphocytes/100 leukocytes in blood by manual count, rate of change in lab result between the most recent and second most recent lab |
| Lab | lymphocytes/100 leukocytes in blood by manual count, most recent lab result |
| Lab | lymphocytes/100 leukocytes in blood by manual count, second most recent lab result |
| Lab | magnesium, rate of change in lab result between the most recent and second most recent lab |
| Lab | magnesium, most recent lab result |
| Lab | magnesium, second most recent lab result |
| Lab | monocytes (#/volume) in blood, rate of change in lab result between the most recent and second most recent lab |
| Lab | monocytes (#/volume) in blood, most recent lab result |
| Lab | monocytes (#/volume) in blood, second most recent lab result |
| Lab | monocytes (#/volume) in blood by automated count, rate of change in lab result between the most recent and second most recent lab |
| Lab | monocytes (#/volume) in blood by automated count, most recent lab result |
| Lab | monocytes (#/volume) in blood by automated count, second most recent lab result |
| Lab | monocytes (#/volume) in blood by manual count, rate of change in lab result between the most recent and second most recent lab |
| Lab | monocytes (#/volume) in blood by manual count, most recent lab result |
| Lab | monocytes (#/volume) in blood by manual count, second most recent lab result |
| Lab | monocytes/100 leukocytes in blood, rate of change in lab result between the most recent and second most recent lab |
| Lab | monocytes/100 leukocytes in blood, most recent lab result |
| Lab | monocytes/100 leukocytes in blood, second most recent lab result |
| Lab | monocytes/100 leukocytes in blood by automated count, rate of change in lab result between the most recent and second most recent lab |
| Lab | monocytes/100 leukocytes in blood by automated count, most recent lab result |
| Lab | monocytes/100 leukocytes in blood by automated count, second most recent lab result |
| Lab | monocytes/100 leukocytes in blood by manual count, rate of change in lab result between the most recent and second most recent lab |
| Lab | monocytes/100 leukocytes in blood by manual count, most recent lab result |
| Lab | monocytes/100 leukocytes in blood by manual count, second most recent lab result |
| Lab | phosphate, rate of change in lab result between the most recent and second most recent lab |
| Lab | phosphate, most recent lab result |
| Lab | phosphate, second most recent lab result |
| Lab | platelets, rate of change in lab result between the most recent and second most recent lab |
| Lab | platelets, most recent lab result |
| Lab | platelets, second most recent lab result |
| Lab | prostatic specific antigen, total, rate of change in lab result between the most recent and second most recent lab |
| Lab | prostate-specific antigen, total, most recent lab result |
| Lab | prostate-specific antigen, total, second most recent lab result |
| Lab | prostate-specific antigen, free in blood/serum/plasma, rate of change in lab result between the most recent and second most recent lab |
| Lab | prostate-specific antigen, free in blood/serum/plasma, most recent lab result |
| Lab | prostate-specific antigen, free in blood/serum/plasma, second most recent lab result |
| Lab | prostate-specific antigen, ratio of free to total, rate of change in lab result between the most recent and second most recent lab |
| Lab | prostate-specific antigen, ratio of free to total, most recent lab result |
| Lab | prostate-specific antigen, ratio of free to total, second most recent lab result |
| Lab | prothrombin time, rate of change in lab result between the most recent and second most recent lab |
| Lab | prothrombin time, most recent lab result |
| Lab | prothrombin time, second most recent lab result |
| Lab | coagulation surface induced, rate of change in lab result between the most recent and second most recent lab |
| Lab | coagulation surface induced, most recent lab result |
| Lab | coagulation surface induced, second most recent lab result |
| Lab | red blood cell count, rate of change in lab result between the most recent and second most recent lab |
| Lab | red blood cell count, most recent lab result |
| Lab | red blood cell count, second most recent lab result |
| Lab | sodium, rate of change in lab result between the most recent and second most recent lab |
| Lab | sodium, most recent lab result |
| Lab | sodium, second most recent lab result |
| Lab | free thyroxine, rate of change in lab result between the most recent and second most recent lab |
| Lab | free thyroxine, most recent lab result |
| Lab | free thyroxine, second most recent lab result |
| Lab | total cholesterol, rate of change in lab result between the most recent and second most recent lab |
| Lab | total cholesterol, most recent lab result |
| Lab | total cholesterol, second most recent lab result |
| Lab | protein, total in blood/serum, rate of change in lab result between the most recent and second most recent lab |
| Lab | protein, total in blood/serum, most recent lab result |
| Lab | protein, total in blood/serum, second most recent lab result |
| Lab | triglycerides, fasting, rate of change in lab result between the most recent and second most recent lab |
| Lab | triglycerides, fasting, most recent lab result |
| Lab | triglycerides, fasting, second most recent lab result |
| Lab | triglycerides, random, rate of change in lab result between the most recent and second most recent lab |
| Lab | triglycerides, random, most recent lab result |
| Lab | triglycerides, random, second most recent lab result |
| Lab | troponin I cardiac quantitative, rate of change in lab result between the most recent and second most recent lab |
| Lab | troponin I cardiac quantitative, most recent lab result |
| Lab | troponin I cardiac quantitative, second most recent lab result |
| Lab | thyroid stimulating hormone, rate of change in lab result between the most recent and second most recent lab |
| Lab | thyroid stimulating hormone, most recent lab result |
| Lab | thyroid stimulating hormone, second most recent lab result |
| Lab | microalbumin or albumin to creatinine ratio in urine, rate of change in lab result between the most recent and second most recent lab |
| Lab | microalbumin or albumin to creatinine ratio in urine, most recent lab result |
| Lab | microalbumin or albumin to creatinine ratio in urine, second most recent lab result |
| Lab | creatinine in urine, rate of change in lab result between the most recent and second most recent lab |
| Lab | creatinine in urine, most recent lab result |
| Lab | creatinine in urine, second most recent lab result |
| Lab | hemoglobin in urine by test strip, rate of change in lab result between the most recent and second most recent lab |
| Lab | hemoglobin in urine by test strip, most recent lab result |
| Lab | hemoglobin in urine by test strip, second most recent lab result |
| Lab | 24-hour microalbumin in urine, rate of change in lab result between the most recent and second most recent lab |
| Lab | 24-hour microalbumin in urine, most recent lab result |
| Lab | 24-hour microalbumin in urine, second most recent lab result |
| Lab | protein to creatinine ratio in urine, rate of change in lab result between the most recent and second most recent lab |
| Lab | protein to creatinine ratio in urine, most recent lab result |
| Lab | protein to creatinine ratio in urine, second most recent lab result |
| Lab | protein in urine, rate of change in lab result between the most recent and second most recent lab |
| Lab | protein in urine, most recent lab result |
| Lab | protein in urine, second most recent lab result |
| Lab | 24-hour urine protein, rate of change in lab result between the most recent and second most recent lab |
| Lab | 24-hour urine protein, most recent lab result |
| Lab | 24-hour urine protein, second most recent lab result |
| Lab | protein in urine by dipstick, qualitative, rate of change in lab result between the most recent and second most recent lab |
| Lab | protein in urine by dipstick, qualitative, most recent lab result |
| Lab | protein in urine by dipstick, qualitative, second most recent lab result |
| Lab | uric acid, rate of change in lab result between the most recent and second most recent lab |
| Lab | uric acid, most recent lab result |
| Lab | uric acid, second most recent lab result |
| Lab | varicella virus result, between 0-6 months prior to index date |
| Lab | varicella virus result, between 7-12 months prior to index date |
| Lab | varicella virus result, between 1-2 years prior to index date |
| Lab | varicella virus result, more than 2 years prior to index date |
| Lab | vitamin B12, rate of change in lab result between the most recent and second most recent lab |
| Lab | vitamin B12, most recent lab result |
| Lab | vitamin B12, second most recent lab result |
| Lab | vitamin D, rate of change in lab result between the most recent and second most recent lab |
| Lab | vitamin D, most recent lab result |
| Lab | vitamin D, second most recent lab result |
| Lab | white blood cell count, total, rate of change in lab result between the most recent and second most recent lab |
| Lab | white blood cell count, total, most recent lab result |
| Lab | white blood cell count, total, second most recent lab result |
| Lab | white blood cell total number, automated count, rate of change in lab result between the most recent and second most recent lab |
| Lab | white blood cell total number, automated count, most recent lab result |
| Lab | white blood cell total number, automated count, second most recent lab result |
| Other | medical record number |
| Other | index date |
| Other | index year |
| Other | year of index date |
| Other | number of days of follow-up |
| Prescriptions | anabolic steroids, between 0-6 months prior to index date |
| Prescriptions | anabolic steroids, between 7-12 months prior to index date |
| Prescriptions | anabolic steroids, between 1-2 years prior to index date |
| Prescriptions | anabolic steroids, more than 2 years prior to index date |
| Prescriptions | anorexia, between 0-6 months prior to index date |
| Prescriptions | anorexia, between 7-12 months prior to index date |
| Prescriptions | anorexia, between 1-2 years prior to index date |
| Prescriptions | anorexia, more than 2 years prior to index date |
| Prescriptions | non-steroidal anti-inflammatory drugs, between 0-6 months prior to index date |
| Prescriptions | non-steroidal anti-inflammatory drugs, between 7-12 months prior to index date |
| Prescriptions | non-steroidal anti-inflammatory drugs, between 1-2 years prior to index date |
| Prescriptions | non-steroidal anti-inflammatory drugs, more than 2 years prior to index date |
| Prescriptions | antirheumatic drug, between 0-6 months prior to index date |
| Prescriptions | antirheumatic drug, between 7-12 months prior to index date |
| Prescriptions | antirheumatic drug, between 1-2 years prior to index date |
| Prescriptions | antirheumatic drug, more than 2 years prior to index date |
| Prescriptions | tumor necrosis factor inhibitor drug, between 0-6 months prior to index date |
| Prescriptions | tumor necrosis factor inhibitor drug, between 7-12 months prior to index date |
| Prescriptions | tumor necrosis factor inhibitor drug, between 1-2 years prior to index date |
| Prescriptions | tumor necrosis factor inhibitor drug, more than 2 years prior to index date |
| Prescriptions | anti-nausea drugs, between 0-6 months prior to index date |
| Prescriptions | anti-nausea drugs, between 7-12 months prior to index date |
| Prescriptions | anti-nausea drugs, between 1-2 years prior to index date |
| Prescriptions | anti-nausea drugs, more than 2 years prior to index date |
| Prescriptions | antivirals, between 0-6 months prior to index date |
| Prescriptions | antivirals, between 7-12 months prior to index date |
| Prescriptions | antivirals, between 1-2 years prior to index date |
| Prescriptions | antivirals, more than 2 years prior to index date |
| Prescriptions | antibiotics, between 0-6 months prior to index date |
| Prescriptions | antibiotics, between 7-12 months prior to index date |
| Prescriptions | antibiotics, between 1-2 years prior to index date |
| Prescriptions | antibiotics, more than 2 years prior to index date |
| Prescriptions | antidepressants, between 0-6 months prior to index date |
| Prescriptions | antidepressants, between 7-12 months prior to index date |
| Prescriptions | antidepressants, between 1-2 years prior to index date |
| Prescriptions | antidepressants, more than 2 years prior to index date |
| Prescriptions | antifungals, between 0-6 months prior to index date |
| Prescriptions | antifungals, between 7-12 months prior to index date |
| Prescriptions | antifungals, between 1-2 years prior to index date |
| Prescriptions | antifungals, more than 2 years prior to index date |
| Prescriptions | antipsychotics, between 0-6 months prior to index date |
| Prescriptions | antipsychotics, between 7-12 months prior to index date |
| Prescriptions | antipsychotics, between 1-2 years prior to index date |
| Prescriptions | antipsychotics, more than 2 years prior to index date |
| Prescriptions | blood thinners, between 0-6 months prior to index date |
| Prescriptions | blood thinners, between 7-12 months prior to index date |
| Prescriptions | blood thinners, between 1-2 years prior to index date |
| Prescriptions | blood thinners, more than 2 years prior to index date |
| Prescriptions | corticosteroids, between 0-6 months prior to index date |
| Prescriptions | corticosteroids, between 7-12 months prior to index date |
| Prescriptions | corticosteroids, between 1-2 years prior to index date |
| Prescriptions | corticosteroids, more than 2 years prior to index date |
| Prescriptions | insulin, between 0-6 months prior to index date |
| Prescriptions | insulin, between 7-12 months prior to index date |
| Prescriptions | insulin, between 1-2 years prior to index date |
| Prescriptions | insulin, more than 2 years prior to index date |
| Prescriptions | diabetes drugs, between 0-6 months prior to index date |
| Prescriptions | diabetes drugs, between 7-12 months prior to index date |
| Prescriptions | diabetes drugs, between 1-2 years prior to index date |
| Prescriptions | diabetes drugs, more than 2 years prior to index date |
| Prescriptions | antihypertensive medications, between 0-6 months prior to index date |
| Prescriptions | antihypertensive medications, between 7-12 months prior to index date |
| Prescriptions | antihypertensive medications, between 1-2 years prior to index date |
| Prescriptions | antihypertensive medications, more than 2 years prior to index date |
| Prescriptions | immunosuppressants, between 0-6 months prior to index date |
| Prescriptions | immunosuppressants, between 7-12 months prior to index date |
| Prescriptions | immunosuppressants, between 1-2 years prior to index date |
| Prescriptions | immunosuppressants, more than 2 years prior to index date |
| Prescriptions | lipid-lowering drugs, between 0-6 months prior to index date |
| Prescriptions | lipid-lowering drugs, between 7-12 months prior to index date |
| Prescriptions | lipid-lowering drugs, between 1-2 years prior to index date |
| Prescriptions | lipid-lowering drugs, more than 2 years prior to index date |
| Prescriptions | metformin, between 0-6 months prior to index date |
| Prescriptions | metformin, between 7-12 months prior to index date |
| Prescriptions | metformin, between 1-2 years prior to index date |
| Prescriptions | metformin, more than 2 years prior to index date |
| Prescriptions | osteoporosis drugs, between 0-6 months prior to index date |
| Prescriptions | osteoporosis drugs, between 7-12 months prior to index date |
| Prescriptions | osteoporosis drugs, between 1-2 years prior to index date |
| Prescriptions | osteoporosis drugs, more than 2 years prior to index date |
| Prescriptions | pain medications, between 0-6 months prior to index date |
| Prescriptions | pain medications, between 7-12 months prior to index date |
| Prescriptions | pain medications, between 1-2 years prior to index date |
| Prescriptions | pain medications, more than 2 years prior to index date |
| Prescriptions | pancreatic enzyme, between 0-6 months prior to index date |
| Prescriptions | pancreatic enzyme, between 7-12 months prior to index date |
| Prescriptions | pancreatic enzyme, between 1-2 years prior to index date |
| Prescriptions | pancreatic enzyme, more than 2 years prior to index date |
| Prescriptions | proton-pump inhibitors, between 0-6 months prior to index date |
| Prescriptions | cumulative dose of proton-pump inhibitors, between 0-6 months prior to index date |
| Prescriptions | proton-pump inhibitors, between 7-12 months prior to index date |
| Prescriptions | cumulative dose of proton-pump inhibitors, between 7-12 months prior to index date |
| Prescriptions | proton-pump inhibitors, between 1-2 years prior to index date |
| Prescriptions | cumulative dose of proton-pump inhibitors, between 1-2 years prior to index date |
| Prescriptions | proton-pump inhibitors, more than 2 years prior to index date |
| Prescriptions | cumulative dose of proton-pump inhibitors, more than 2 years prior to index date |
| Prescriptions | ulcer drugs, between 0-6 months prior to index date |
| Prescriptions | ulcer drugs, between 7-12 months prior to index date |
| Prescriptions | ulcer drugs, between 1-2 years prior to index date |
| Prescriptions | ulcer drugs, more than 2 years prior to index date |
| Procedures | abdominal and chest CT, between 0-6 months prior to index date |
| Procedures | abdominal and chest CT, between 7-12 months prior to index date |
| Procedures | abdominal and chest CT, between 1-2 years prior to index date |
| Procedures | abdominal and chest CT, more than 2 years prior to index date |
| Procedures | abdominal and chest MRI, between 0-6 months prior to index date |
| Procedures | abdominal and chest MRI, between 7-12 months prior to index date |
| Procedures | abdominal and chest MRI, between 1-2 years prior to index date |
| Procedures | abdominal and chest MRI, more than 2 years prior to index date |
| Procedures | abdominal ultrasound, between 0-6 months prior to index date |
| Procedures | abdominal ultrasound, between 7-12 months prior to index date |
| Procedures | abdominal ultrasound, between 1-2 years prior to index date |
| Procedures | abdominal ultrasound, more than 2 years prior to index date |
| Procedures | barium-meal photofluorography, between 0-6 months prior to index date |
| Procedures | barium-meal photofluorography, between 7-12 months prior to index date |
| Procedures | barium-meal photofluorography, between 1-2 years prior to index date |
| Procedures | barium-meal photofluorography, more than 2 years prior to index date |
| Procedures | colonoscopy, between 0-6 months prior to index date |
| Procedures | colonoscopy, between 7-12 months prior to index date |
| Procedures | colonoscopy, between 1-2 years prior to index date |
| Procedures | colonoscopy, more than 2 years prior to index date |
| Procedures | sigmoidoscopy, between 0-6 months prior to index date |
| Procedures | sigmoidoscopy, between 7-12 months prior to index date |
| Procedures | sigmoidoscopy, between 1-2 years prior to index date |
| Procedures | sigmoidoscopy, more than 2 years prior to index date |
| Procedures | surgical procedures on the abdomen, peritoneum, and omentum, between 0-6 months prior to index date |
| Procedures | surgical procedures on the abdomen, peritoneum, and omentum, between 7-12 months prior to index date |
| Procedures | surgical procedures on the abdomen, peritoneum, and omentum, between 1-2 years prior to index date |
| Procedures | surgical procedures on the abdomen, peritoneum, and omentum, more than 2 years prior to index date |
| Procedures | surgical procedures on the anus, between 0-6 months prior to index date |
| Procedures | surgical procedures on the anus, between 7-12 months prior to index date |
| Procedures | surgical procedures on the anus, between 1-2 years prior to index date |
| Procedures | surgical procedures on the anus, more than 2 years prior to index date |
| Procedures | surgical procedures on the appendix, between 0-6 months prior to index date |
| Procedures | surgical procedures on the appendix, between 7-12 months prior to index date |
| Procedures | surgical procedures on the appendix, between 1-2 years prior to index date |
| Procedures | surgical procedures on the appendix, more than 2 years prior to index date |
| Procedures | surgical procedures on the biliary tract, between 0-6 months prior to index date |
| Procedures | surgical procedures on the biliary tract, between 7-12 months prior to index date |
| Procedures | surgical procedures on the biliary tract, between 1-2 years prior to index date |
| Procedures | surgical procedures on the biliary tract, more than 2 years prior to index date |
| Procedures | surgical procedures on Meckel's diverticulum and the mesentery, between 0-6 months prior to index date |
| Procedures | surgical procedures on Meckel's diverticulum and the mesentery, between 7-12 months prior to index date |
| Procedures | surgical procedures on Meckel's diverticulum and the mesentery, between 1-2 years prior to index date |
| Procedures | surgical procedures on Meckel's diverticulum and the mesentery, more than 2 years prior to index date |
| Procedures | surgical procedures on the esophagus, between 0-6 months prior to index date |
| Procedures | surgical procedures on the esophagus, between 7-12 months prior to index date |
| Procedures | surgical procedures on the esophagus, between 1-2 years prior to index date |
| Procedures | surgical procedures on the esophagus, more than 2 years prior to index date |
| Procedures | surgical procedures on the intestines (except rectum), between 0-6 months prior to index date |
| Procedures | surgical procedures on the intestines (except rectum), between 7-12 months prior to index date |
| Procedures | surgical procedures on the intestines (except rectum), between 1-2 years prior to index date |
| Procedures | surgical procedures on the intestines (except rectum), more than 2 years prior to index date |
| Procedures | surgical procedures on the liver, between 0-6 months prior to index date |
| Procedures | surgical procedures on the liver, between 7-12 months prior to index date |
| Procedures | surgical procedures on the liver, between 1-2 years prior to index date |
| Procedures | surgical procedures on the liver, more than 2 years prior to index date |
| Procedures | surgical procedures on the pancreas, between 0-6 months prior to index date |
| Procedures | surgical procedures on the pancreas, between 7-12 months prior to index date |
| Procedures | surgical procedures on the pancreas, between 1-2 years prior to index date |
| Procedures | surgical procedures on the pancreas, more than 2 years prior to index date |
| Procedures | surgical procedures on the rectum, between 0-6 months prior to index date |
| Procedures | surgical procedures on the rectum, between 7-12 months prior to index date |
| Procedures | surgical procedures on the rectum, between 1-2 years prior to index date |
| Procedures | surgical procedures on the rectum, more than 2 years prior to index date |
| Procedures | surgical procedures on the stomach, between 0-6 months prior to index date |
| Procedures | surgical procedures on the stomach, between 7-12 months prior to index date |
| Procedures | surgical procedures on the stomach, between 1-2 years prior to index date |
| Procedures | surgical procedures on the stomach, more than 2 years prior to index date |
| Procedures | organ transplantation, between 0-6 months prior to index date |
| Procedures | organ transplantation, between 7-12 months prior to index date |
| Procedures | organ transplantation, between 1-2 years prior to index date |
| Procedures | organ transplantation, more than 2 years prior to index date |
| Procedures | upper GI endoscopy, between 0-6 months prior to index date |
| Procedures | upper GI endoscopy, between 7-12 months prior to index date |
| Procedures | upper GI endoscopy, between 1-2 years prior to index date |
| Procedures | upper GI endoscopy, more than 2 years prior to index date |
| Symptoms | abdominal pain, between 0-6 months prior to index date |
| Symptoms | abdominal pain, between 7-12 months prior to index date |
| Symptoms | abdominal pain, between 1-2 years prior to index date |
| Symptoms | abdominal pain, more than 2 years prior to index date |
| Symptoms | anorexia, between 0-6 months prior to index date |
| Symptoms | anorexia, between 7-12 months prior to index date |
| Symptoms | anorexia, between 1-2 years prior to index date |
| Symptoms | anorexia, more than 2 years prior to index date |
| Symptoms | back pain, between 0-6 months prior to index date |
| Symptoms | back pain, between 7-12 months prior to index date |
| Symptoms | back pain, between 1-2 years prior to index date |
| Symptoms | back pain, more than 2 years prior to index date |
| Symptoms | change in bowel habit, between 0-6 months prior to index date |
| Symptoms | change in bowel habit, between 7-12 months prior to index date |
| Symptoms | change in bowel habit, between 1-2 years prior to index date |
| Symptoms | change in bowel habit, more than 2 years prior to index date |
| Symptoms | chest pain, between 0-6 months prior to index date |
| Symptoms | chest pain, between 7-12 months prior to index date |
| Symptoms | chest pain, between 1-2 years prior to index date |
| Symptoms | chest pain, more than 2 years prior to index date |
| Symptoms | constipation, between 0-6 months prior to index date |
| Symptoms | constipation, between 7-12 months prior to index date |
| Symptoms | constipation, between 1-2 years prior to index date |
| Symptoms | constipation, more than 2 years prior to index date |
| Symptoms | diarrhea, between 0-6 months prior to index date |
| Symptoms | diarrhea, between 7-12 months prior to index date |
| Symptoms | diarrhea, between 1-2 years prior to index date |
| Symptoms | diarrhea, more than 2 years prior to index date |
| Symptoms | itching, between 0-6 months prior to index date |
| Symptoms | itching, between 7-12 months prior to index date |
| Symptoms | itching, between 1-2 years prior to index date |
| Symptoms | itching, more than 2 years prior to index date |
| Symptoms | malaise or fatigue, between 0-6 months prior to index date |
| Symptoms | malaise or fatigue, between 7-12 months prior to index date |
| Symptoms | malaise or fatigue, between 1-2 years prior to index date |
| Symptoms | malaise or fatigue, more than 2 years prior to index date |
| Symptoms | melena, between 0-6 months prior to index date |
| Symptoms | melena, between 7-12 months prior to index date |
| Symptoms | melena, between 1-2 years prior to index date |
| Symptoms | melena, more than 2 years prior to index date |
| Symptoms | nausea or vomiting, between 0-6 months prior to index date |
| Symptoms | nausea or vomiting, between 7-12 months prior to index date |
| Symptoms | nausea or vomiting, between 1-2 years prior to index date |
| Symptoms | nausea or vomiting, more than 2 years prior to index date |
| Symptoms | swollen lymph node, between 0-6 months prior to index date |
| Symptoms | swollen lymph node, between 7-12 months prior to index date |
| Symptoms | swollen lymph node, between 1-2 years prior to index date |
| Symptoms | swollen lymph node, more than 2 years prior to index date |
| Utilization | total emergency department utilization, between 0-6 months prior to index date |
| Utilization | total emergency department utilization, between 7-12 months prior to index date |
| Utilization | total emergency department utilization, between 1-2 years prior to index date |
| Utilization | total emergency department utilization, more than 2 years prior to index date |
| Utilization | total inpatient utilization, between 0-6 months prior to index date |
| Utilization | total inpatient utilization, between 7-12 months prior to index date |
| Utilization | total inpatient utilization, between 1-2 years prior to index date |
| Utilization | total inpatient utilization, more than 2 years prior to index date |
| Utilization | total outpatient utilization, between 0-6 months prior to index date |
| Utilization | total outpatient utilization, between 7-12 months prior to index date |
| Utilization | total outpatient utilization, between 1-2 years prior to index date |
| Utilization | total outpatient utilization, more than 2 years prior to index date |
| Utilization | total urgent care utilization, between 0-6 months prior to index date |
| Utilization | total urgent care utilization, between 7-12 months prior to index date |
| Utilization | total urgent care utilization, between 1-2 years prior to index date |
| Utilization | total urgent care utilization, more than 2 years prior to index date |

**eTable 3: Definitions of weight/laboratory value changes and symptom-related features.**

| **Features Category** | **Relevant features** |
| --- | --- |
| Lab measures and changes | - Most recent value: the value on the index date or closest to and within 6 months prior to the index date. - Second most recent value: a prior value that was within 9 to 15 months prior to the most recent value. - Absolute change: the difference between the most recent value and the second most recent value. - Change rate = (the most recent value – second recent value) / (the most recent value’s measurement date – the second recent value’s measurement date). |
| Weight measures and change | - Most recent value: the value on the index date or closest to and within 12 months prior to the index date. - Second most recent value: a prior value that was older than most recent value and was not on the same day as the most recent value, and within 12 months prior to the index date. - Absolute change: the difference between the most recent value and the second most recent value. - Change rate = (the most recent value – second recent value) / (the most recent value’s measurement date – the second recent value’s measurement date). |
| Symptoms | Four indicators were created reflecting the status within each of the following time windows:   - 0-6 months prior to index date, - 7-12 months prior to index date, - 1-2 years prior to index date, - More than 2 years prior to index date.   The following features combining time windows were created. For example, let X1, X2, X3 and X4 be the four indicators indicating the presence/absence of a symptom-based feature (e.g. constipation) in the four time windows described above, the follow three additional features were generated.  X12=X1 or X2  X123=X1 or X2 or X3  X1234=X1 or X2 or X3 or X4 |

**eTable 4. Feature selection process and preselected potential features for the main cohort and early detection cohort.**

***Main cohort:***

***First round: started with 511 potential features and ended with 63 potential features.*** A feature was selected if its minimal depth ≤ 5.2 in at least one out of 10 imputed datasets.

***Second round: started with 63 potential features and ended with 45 potential features.*** A feature was selected if its minimal depth ≤ 5.0 in at least one of 10 imputed datasets.

***Third round: started with 45 potential features and ended with 29 potential features.*** A feature was selected if its minimal depth ≤ 4.8 in at least one of 10 imputed datasets.

***Early Detection cohort:***

***First round: started with 511 potential features and ended with 38 potential features.*** A feature was selected if its minimal average depth ≤ 5.2 in at least one out of 10 imputed datasets.

***Second round: started with 38 potential features and ended with 37 potential features.*** A feature was selected if its minimal average depth ≤ 5.0 in at least one of 10 imputed datasets.

***Third round: started with 37 potential features and ended with 32 potential features.*** A feature was selected if its minimal average depth ≤ 4.8 in at least one of 10 imputed datasets.

| Pre-Selected Features | Main Cohort | Early Detection Cohort |
| --- | --- | --- |
| age at index date | x | x |
| alanine aminotransferase, rate of change in lab result between the most recent and second most recent lab | x | x |
| alanine aminotransferase, difference in results between the most recent and second most recent labs | x | x |
| acute pancreatitis, between 0-6 months prior to index date | x | x |
| weight change per day, calculated from the two farthest values within the year prior to index date | x | x |
| hemoglobin A1c, most recent lab result | x | x |
| alanine aminotransferase, most recent lab result | x | x |
| surgical procedures on the esophagus, between 0-6 months prior to index date | x | x |
| hemoglobin A1c, rate of change in lab result between the most recent and second most recent lab | x | x |
| abdominal pain, between 0-6 months prior to index date | x |  |
| chronic pancreatitis, between 0-2 years prior to index date | x |  |
| BMI closest and prior to index date | x |  |
| hemoglobin A1c, difference in results between the most recent and second most recent labs | x | x |
| red blood cell count, most recent lab result | x | x |
| total cholesterol, most recent lab result | x |  |
| benign pancreatic disorders, between 0-6 months prior to index date | x | x |
| hematocrit in blood, most recent lab result | x | x |
| historical infection of Helicobacter pylori result, between 0-6 months prior to index date | x |  |
| chronic pancreatitis, between 0-6 months prior to index date | x | x |
| constipation, between 0-6 months prior to index date | x |  |
| melena, between 0-6 months prior to index date | x |  |
| hemoglobin, rate of change in lab result between the most recent and second most recent lab | x |  |
| hemoglobin, most recent lab result | x |  |
| hematocrit in blood, difference in results between the most recent and second most recent labs | x | x |
| pancreatic enzyme, between 0-6 months prior to index date | x | x |
| red blood cell count, rate of change in lab result between the most recent and second most recent lab | x |  |
| platelets, rate of change in lab result between the most recent and second most recent lab | x |  |
| chronic pancreatitis, between 0-12 months prior to index date | x |  |
| chronic pancreatitis, any time prior to index date | x |  |
| hemoglobin A1c, second most recent lab result |  | x |
| red blood cell count, second most recent lab result |  | x |
| sodium, rate of change in lab result between the most recent and second most recent lab |  | x |
| Charlson comorbidity index within one year prior to index date |  | x |
| type of diabetes mellitus, between 0-12 months prior to index date |  | x |
| type of diabetes mellitus, any time prior to index date |  | x |
| benign pancreatic disorders,between 0-2 years prior to index date |  | x |
| benign pancreatic disorders, any time prior to index date |  | x |
| chronic pancreatitis, between 0-12 months prior to index date |  | x |
| chronic pancreatitis, between 0-2 years prior to index date |  | x |
| chronic pancreatitis, any time prior to index date |  | x |
| chronic pancreatitis, between 1-2 years prior to index date |  | x |
| biliary tract disease, between 0-6 months prior to index date |  | x |
| hypertension, more than 2 years prior to index date |  | x |
| total outpatient utilization, more than 2 years prior to index date |  | x |
| weight closest and prior to index date |  | x |

**eTable 5. Distribution of American Joint Committee on Cancer (AJCC) stage in patients with PDAC.**

|  | KP cohort | | VA Cohort | |
| --- | --- | --- | --- | --- |
| N | 1,492 | | 2,018 | |
|  | Count | Percent | Count | Percent |
| Stage I | 96 | 6.4% | 103 | 5.1% |
| Stage II | 377 | 25.3% | 282 | 14.0% |
| Stage III | 122 | 8.2% | 106 | 5.3% |
| Stage IV | 678 | 45.4% | 551 | 27.3% |
| Unknown/Missing | 219 | 14.7% | 976 | 48.4% |

Abbreviations: KPSC, Kaiser Permanente Southern California; VA, Veterans Affairs.

**eTable 6. Number and size of KPSC training, KPSC internal validation and VA external testing datasets.**

|  | **Main Cohort** | | **Early Detection Cohort** | |
| --- | --- | --- | --- | --- |
| **Datasets** | **Number** | **Size** | **Number** | **Size** |
| KPSC training | 50 | 1,441,546 | 50 | 1,379,903 |
| KPSC internal validation | 50 | 360,386 | 50 | 344,976 |
| KPSC internal validation; restricted to patients with complete 18 months follow up or those who developed PDAC in 18 months | 50 | 249,736 | 50 | 235,041 |
| VA external testing – recalibrated model | 10 | 2,690,895 | 10 | 2,633,112 |
| VA external testing – recalibrated model; restricted to patients with complete 18 months follow up or those who developed PDAC in 18 months | 10 | 2,122,288 | 10 | 2,121,832 |
| VA external testing – directly applied model | 5 | 2,690,895 | Did not perform | Did not perform |

Abbreviations: KPSC, Kaiser Permanente Southern California; VA, Veterans Affairs.

**eTable 7. Hyperparameter setup for both feature pre-selection and model development for the main and early detection cohorts.**

|  | **Main Cohort** | | | | |
| --- | --- | --- | --- | --- | --- |
|  | **ntree** | **depth*** | **mtry** | **nodesize** | **bagging percentage** |
| First round of feature preselection | 50 | 6 | 23 | 150 | 0.632 |
| Second round of feature preselection | 50 | 6 | 8 | 150 | 0.632 |
| Third round of feature preselection | 50 | 6 | 6 | 150 | 0.632 |
| Model development | 20 | Searched in the range of 6-10. 6 were selected. | Number of features | 150 | 0.632 |
|  | Early Detection Cohort | | | | |
|  | ntree | depth* | mtry | nodesize | bagging percentage |
| First round of feature preselection | 50 | 6 | 23 | 150 | 0.632 |
| Second round of feature preselection | 50 | 6 | 7 | 150 | 0.632 |
| Third round of feature preselection | 50 | 6 | 7 | 150 | 0.632 |
| Model development | 20 | 6 | Number of features | 15** | 0.632 |

All hyperparameters were fixed unless otherwise stated.

* This number does not include terminal nodes.

** The default parameter was applied.

**eTable 8. Predictors selected by at least 5 out of the 50 models based on the 50 training samples.**

| **Main Cohort** | | **Early Detection Cohort** | |
| --- | --- | --- | --- |
| **Selected Predictor (ranked from highest to lowest)** | **Frequency of appearance in 50 training samples**  **n (%)** | **Selected Predictor (ranked from highest to lowest)** | **Frequency of appearance in 50 training samples**  **n (%)** |
| Age* | NA | Age* | NA |
| Abdominal pain | 50 (100%) | Rate of ALT change | 36 (72%) |
| Weight change in 1 year | 34 (68%) | Weight change per day | 36 (72%) |
| HgA1c | 30 (60%) | HgA1c | 30 (60%) |
| Rate of ALT change | 28 (56%) | ALT change | 16 (32%) |
| ALT change | 23 (46%) | Rate of HgA1c change | 15 (30%) |
| Rate of HgA1c change | 20 (40%) | HgA1c change | 9 (18%) |
|  |  | Esophagus procedure in past 6 m | 5 (10%) |
|  |  | Any history of chronic pancreatitis | 5 (10%) |

Abbreviations: ALT, alanine transaminase; HgA1c, hemoglobin A1c; NA, not applicable.

*Forced into the models.
