## Supplementary figures and images for "Prediction Model for Detection of Sporadic Pancreatic Cancer (PRO-TECT) in a Population-Based Cohort Using Machine Learning and Further Validation in a Prospective Study"

### eFigure 1

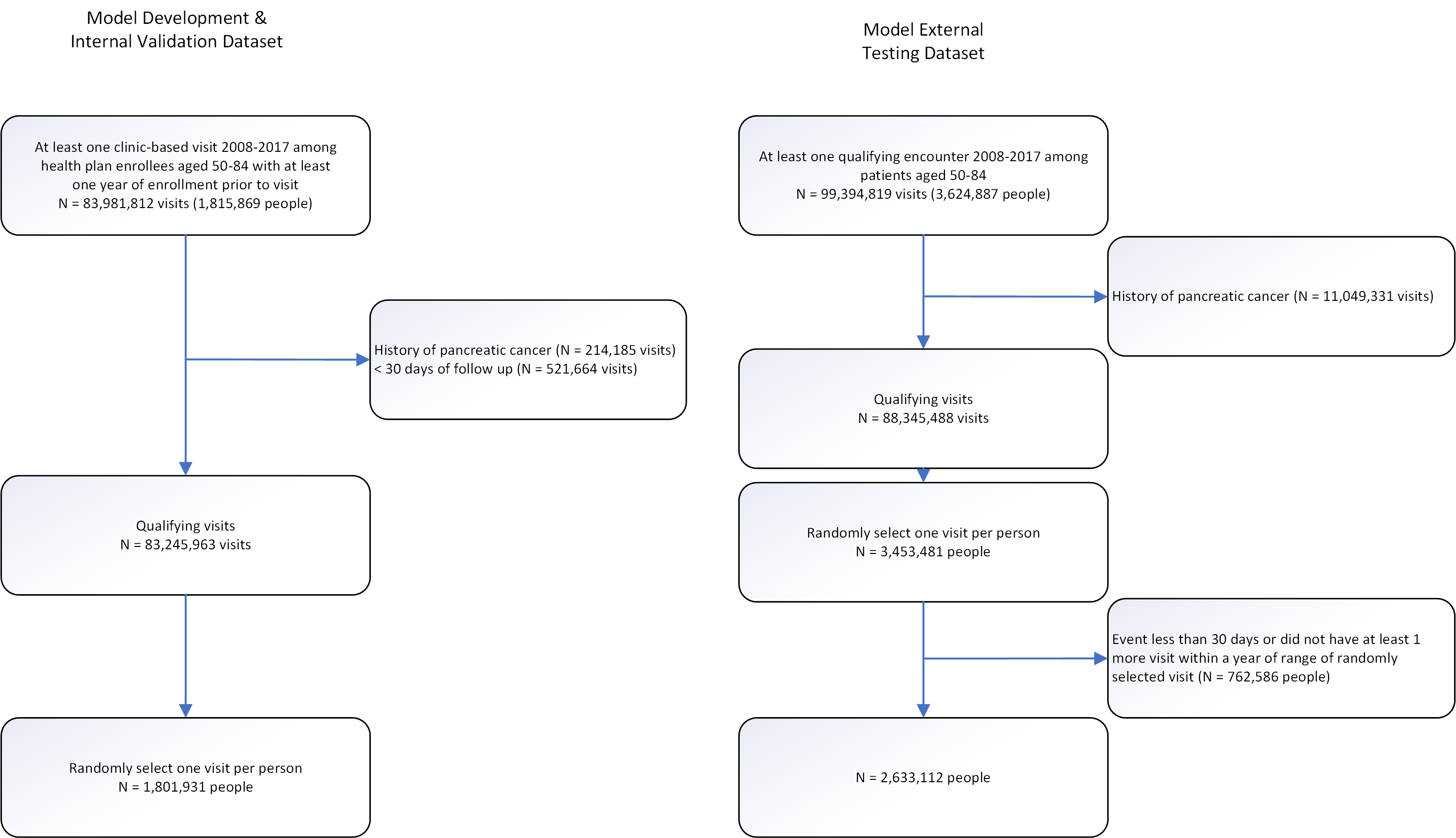

### eFigure 2

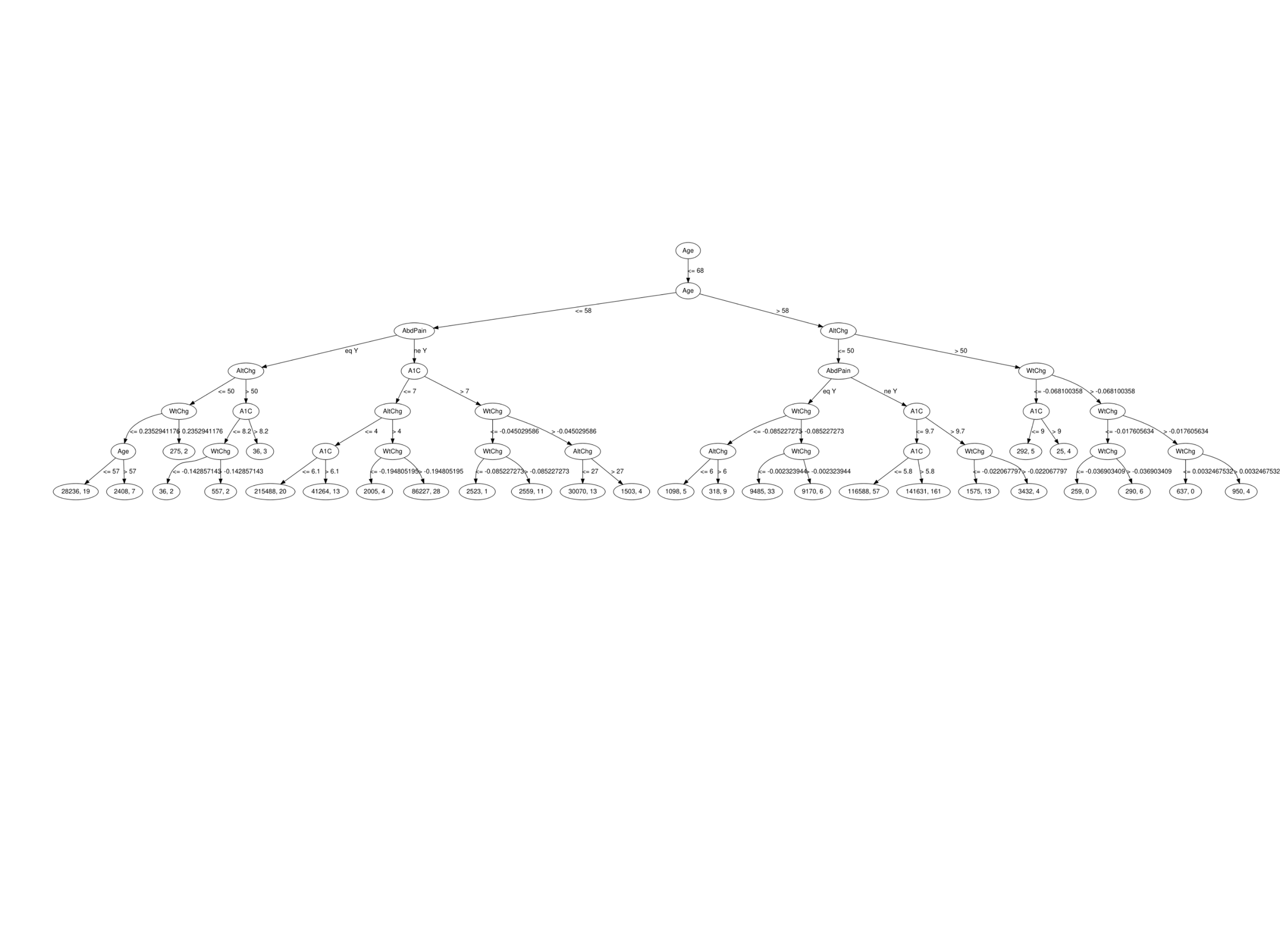

### eFigure 3

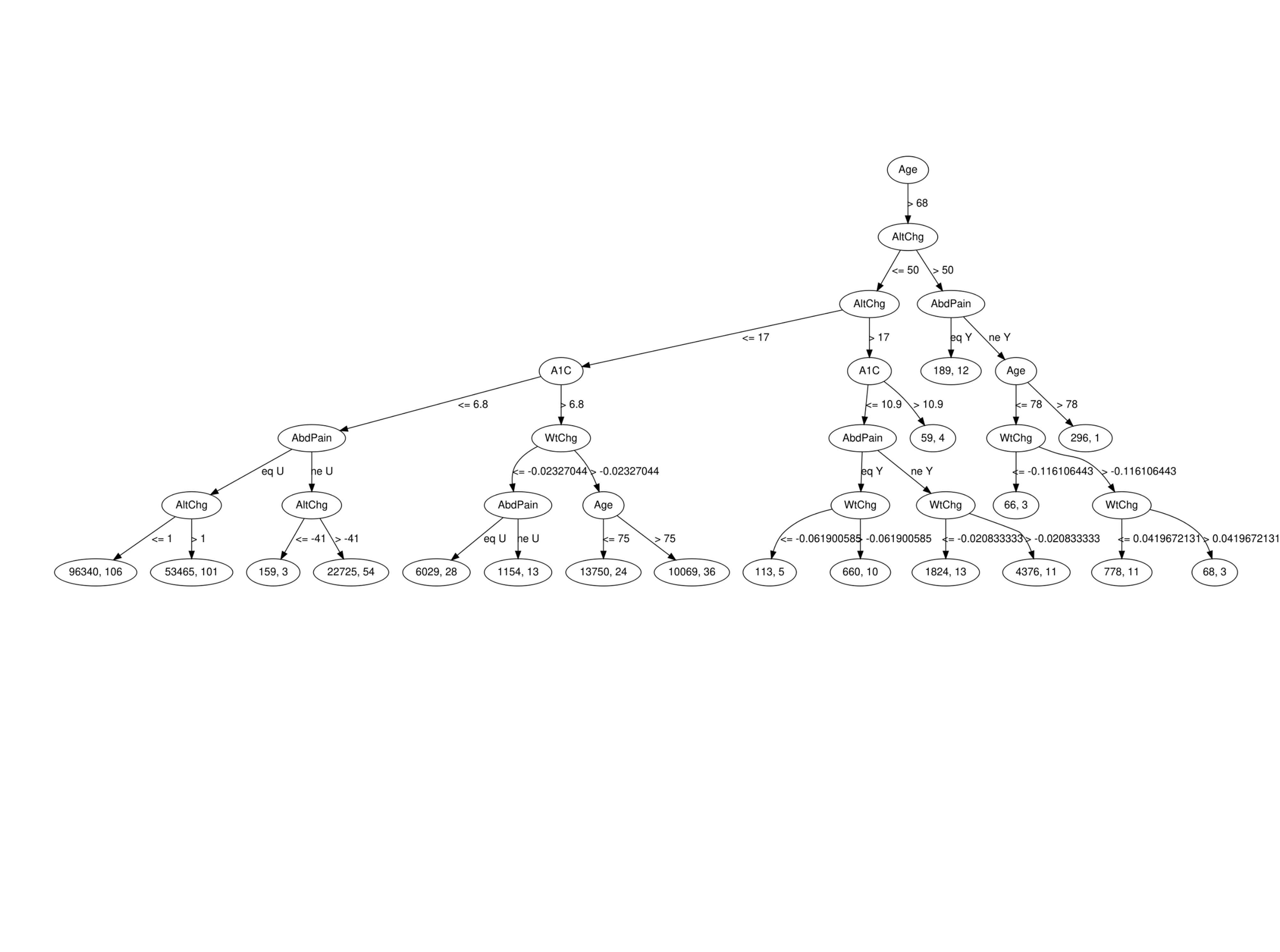
